# Exome sequencing in bipolar disorder reveals shared risk gene *AKAP11* with schizophrenia

**DOI:** 10.1101/2021.03.09.21252930

**Authors:** Duncan S Palmer, Daniel P Howrigan, Sinéad B Chapman, Rolf Adolfsson, Nick Bass, Douglas Blackwood, Marco PM Boks, Chia-Yen Chen, Claire Churchhouse, Aiden P Corvin, Nicholas Craddock, David Curtis, Arianna Di Florio, Faith Dickerson, Fernando S Goes, Xiaoming Jia, Ian Jones, Lisa Jones, Lina Jonsson, Rene S Kahn, Mikael Landén, Adam Locke, Andrew McIntosh, Andrew McQuillin, Derek W Morris, Michael C O’Donovan, Roel A Ophoff, Michael J Owen, Nancy Pedersen, Danielle Posthuma, Andreas Reif, Neil Risch, Catherine Schaefer, Laura Scott, Tarjinder Singh, Jordan W Smoller, Matthew Solomonson, David St. Clair, Eli A Stahl, Annabel Vreeker, James Walters, Weiqing Wang, Nicholas A Watts, Robert Yolken, Peter Zandi, Benjamin M Neale

## Abstract

Here we report results from the Bipolar Exome (BipEx) collaboration analysis of whole exome sequencing of 13,933 individuals diagnosed with bipolar disorder (BD), matched with 14,422 controls. We find an excess of ultra-rare protein-truncating variants (PTVs) in BD patients among genes under strong evolutionary constraint, a signal evident in both major BD subtypes, bipolar 1 disorder (BD1) and bipolar 2 disorder (BD2). We also find an excess of ultra-rare PTVs within genes implicated from a recent schizophrenia exome meta-analysis (SCHEMA; 24,248 SCZ cases and 97,322 controls) and among binding targets of CHD8. Genes implicated from GWAS of BD, however, are not significantly enriched for ultra-rare PTVs. Combining BD gene-level results with SCHEMA, *AKAP11* emerges as a definitive risk gene (ultra-rare PTVs seen in 33 cases and 13 controls, OR = 7.06, *P* = 2.83 × 10^−9^). At the protein level, AKAP-11 is known to interact with GSK3B, the hypothesized mechanism of action for lithium, one of the few treatments for BD. Overall, our results lend further support to the polygenic basis of BD and demonstrate a role for rare coding variation as a significant risk factor in BD onset.

## Introduction

Bipolar disorder (BD) is a heritable neuropsychiatric disorder characterized by episodes of (hypo-)mania and episodes of depression. Across the lifetime, BD has a prevalence between 1-2% of the population, often with onset in early adulthood. Bipolar disorder is a chronic condition that affects individuals across their lifespan and is a significant source of disease burden worldwide (*1*). Meta-analysis of 24 twin studies estimated broad heritability of BD around 67% (*2*), while recent molecular genetic analyses estimated the additive heritable component from common SNPs (MAF > 1%) between 17 and 23% (*3*). This difference between twin heritability estimates of BD and additive heritability from common SNPs indicates that a large fraction of genetic risk is still undiscovered. One potential source may come from rare, often deleterious, genetic variants of more recent origin.

Rare variation, particularly copy number variants, have been shown to influence risk for BD, albeit to a weaker degree than other neuropsychiatric illnesses such as schizophrenia and autism spectrum disorders (ASDs) (*4*). Similarly, previous studies showed some evidence for the role of rare PTVs in BD risk, but with a more modest effect size compared to ASDs and schizophrenia (*5*). The extent that rare variation plays a role in BD susceptibility can be inferred by measuring the degree of natural selection acting on individuals with BD. Specifically, negative selection on BD causes alleles with high penetrance for BD risk to be kept at low frequency in the population (*6, 7*). Evidence for negative selection on BD can be seen in the significantly lower reproductive rate of both males (0.75 to 1) and females (0.85 to 1) with BD compared to their unaffected siblings in a large Swedish birth cohort (*8*). The reproductive rate observed in BD, however, is substantially higher than individuals with schizophrenia (0.23 for males, 0.47 for females) or autism (0.25 for males, 0.48 for females), suggesting that the role of rare variation is likely to be smaller in magnitude, as selection is not acting as strongly on BD in aggregate. Nevertheless, the interrogation of rare variation in BD patients will be pivotal in the discovery of variants with high penetrance for BD risk.

Within BD, two clinical subtype classifications are recognized: bipolar I disorder (BD1) and bipolar II disorder (BD2; APA DSM-IV (*9*); WHO ICD-10 (*10*)). BD1 diagnosis includes recurring manic and depressive episodes, with manic episodes often including psychosis symptoms. In contrast, a BD2 diagnosis requires at least one depressive episode and one hypomanic (but not manic) episode across the lifetime. In addition to the primary BD1/BD2 criteria, individuals presenting with both mood symptoms of BD and psychotic symptoms similar to schizophrenia outside of mood episodes are often diagnosed with schizoaffective disorder (SAD)-bipolar subtype. Despite the distinct diagnostic categories, genetic susceptibility for BD from common SNPs has shown strong overlap with schizophrenia (genetic correlation *r*_g_ = 0.70) and major depressive disorder (MDD) (*r*_g_ = 0.35), with BD1 showing preferential overlap with schizophrenia and BD2 with MDD, reflecting a broad continuum of genetic influence on psychosis and mood disturbance (*3*).

To date, GWAS meta-analysis of common SNPs have now identified 64 independent loci that contribute to BD susceptibility, implicating genes encoding ion channels, neurotransmitter transporters, and synaptic and calcium signalling pathways (*3, 11*). Evidence of rare variation on BD risk, however, remains inconclusive as sample sizes are substantially smaller than GWAS. Analysis of large rare copy number variants (MAF < 1%) in 6,353 BD cases found CNV enrichment among SAD over both controls and other BD diagnoses, suggesting that increased risk among detectable rare CNVs is restricted to individuals with psychotic symptoms (*4*). Analysis of whole exome and genome sequencing of both pedigree and case-control to date have shown only nominal enrichment among individual genes and candidate gene sets (*12*–*15*), with none surpassing exome-wide significance.

Here, we report results from the Bipolar Exome (BipEx) collaboration, the largest whole-exome study of BD to date, comprising 13,933 BD cases and 14,422 controls following aggregation, sequencing, and quality control.

## Results

### Description of exome sequencing data generation, sample cohorts and quality control

We combined bipolar case-control whole exome sequencing data from 13 sample collections in 6 countries. The aggregated dataset consists of 33,699 individuals, 16,486 of which have been diagnosed with bipolar disorder, and 17,213 with no known psychiatric diagnosis (See Table S1 and supplementary materials: sample collections, for a full breakdown by cohort and subtype, and subtype definitions). All of the sample collections have been previously genotyped for common variant analyses (*3*). However, this is the first time that exome-sequencing has been performed and jointly analysed. All exome sequence data was generated using the same library preparation, sequencing platform, and joint calling pipeline: exome sequencing of the full sample set was performed between July 2017 and September 2018 using Illumina Nextera sample preparation and HiSeqX sequencing. Samples were then jointly processed and run through variant calling using the Genome Analysis ToolKit (GATK), (supplementary materials: sequence data production). Following sequencing and joint calling, we ran a series of quality control steps to filter out low quality variants (Table S2) and samples (Table S3), restricting to unrelated individuals of broad continental European ancestry (supplementary materials: exome quality control, Figures S1-5). The analysis-ready high-quality dataset consisting of 13,933 bipolar cases and 14,422 controls is summarised in Table S4. Breaking down by subtype, the curated dataset consists of 8,238 BD1, 3,446 BD2, 1,288 BDNOS, 961 BD without a finer diagnosis (together encompassing the 13,933 BD), and 277 SAD. Throughout our analyses, we exclude individuals diagnosed with SAD in order to obtain a more BD specific collection of results and guard against signals more attributable to schizophrenia being pulled into any reported associations.

### Significant contribution of rare damaging protein truncating variation to bipolar risk

To test whether bipolar cases carry an excess of damaging coding variants, we analyzed exome-wide burden relative to controls using a logistic regression model controlling for principal components, sex, and overall coding burden (supplementary materials: exome-wide burden analyses). Drawing from previous exome sequencing studies of psychiatric disease (*14, 16, 17*), we restricted our attention to variants with minor allele count (MAC) ≤ 5 across the entirety of the dataset, corresponding to MAF ≤ 0.01%. We annotated variants using the Ensembl Variant Effect Predictor (VEP) (*18*) version 95 with the loftee plugin, and assigned variants to classes of variation. We defined three putatively damaging classes of coding variation: protein-truncating variants (PTVs), missense variants with MPC > 2, and damaging missense variants (missense variants annotated as ‘probably damaging’ in PolyPhen and ‘deleterious’ in SIFT). We further defined two annotations which we hypothesised to be likely benign: other missense (the remaining missense variants), and synonymous variants (see supplementary materials: variant annotation and Table S5 for full details). Following this initial restriction we observed nominally significant enrichment of damaging missense variation in BD cases and BD2 cases over controls (OR = 1.01, *P =* 0.024 and OR = 1.02, *P =* 0.0086 respectively); Figure 1B,C, but not for the other *a priori* damaging classes of variation (missense MPC > 2, and PTV). However, stepwise filtering of rare PTVs to those not in the non-psychiatric portion of the Genome Aggregation Database (gnomAD), hereafter referred to as ‘ultra-rare variants’, and then in constrained genes (defined as *p*LI *≥* 0.9), shows that case-control PTV enrichment is present once we filter to high *p*LI genes, a finding in line with schizophrenia exomes (*19*); Figure 1B,C. This enrichment is consistent among both BD1 and BD2 subtypes (Figure 1A). While the magnitude of PTV enrichment in BD (OR = 1.11, *P =* 5.0 × 10^−5^) is considerably lower than the latest PTV enrichment in schizophrenia (OR = 1.26; (*19*)), this difference is in line with the increased selective pressure estimated from higher reproductive rates in BD affected siblings relative to those seen in schizophrenia affected siblings (*8*).

**Figure 1:**
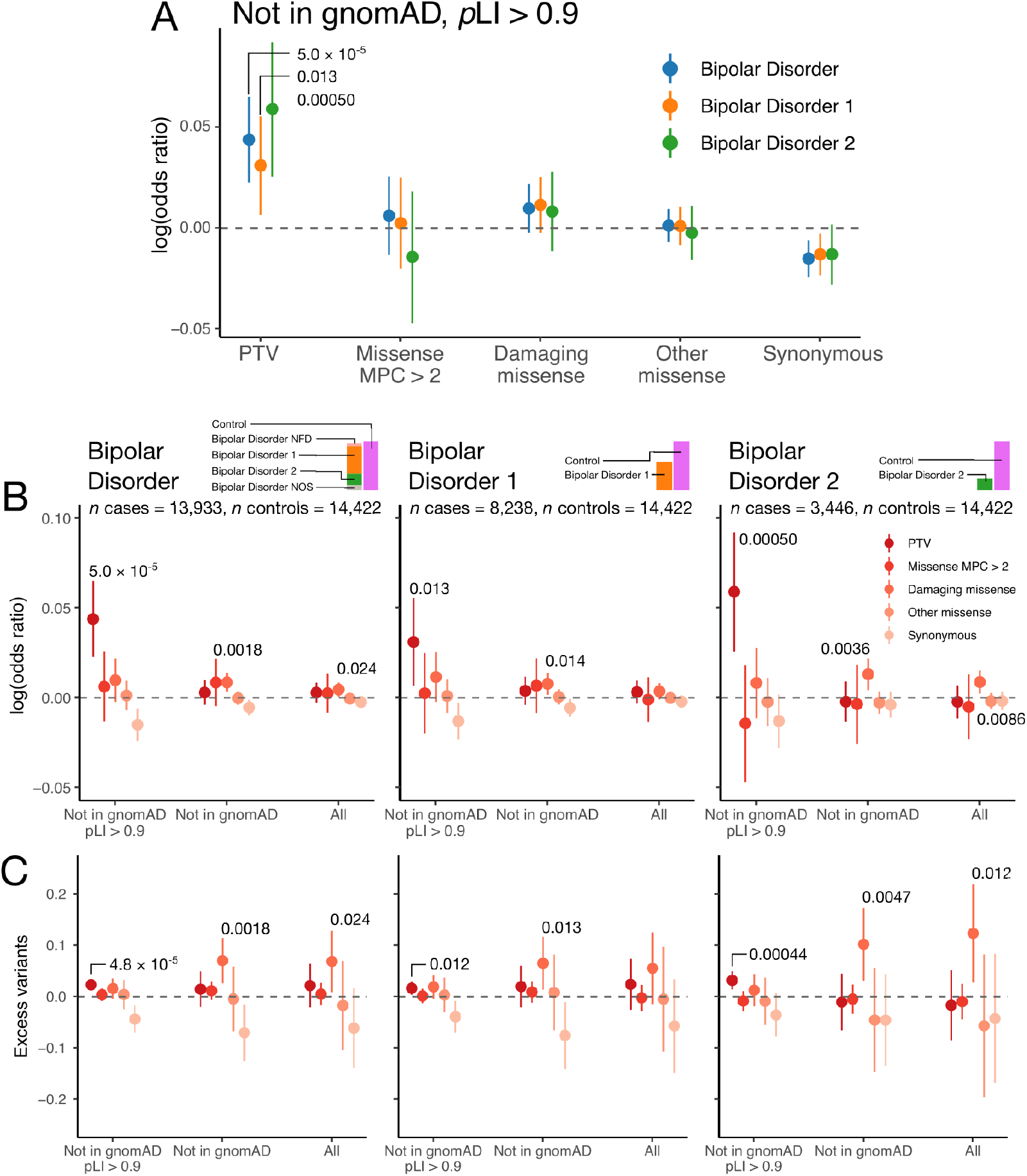
Case-control enrichment of ultra-rare variants, split by case status and consequence category. Panel A displays enrichment in cases over controls in case subsets, according to the legend. In panels B and C, we display case-control enrichment and excess case rare variant burden increasingly *a priori* damaging variant subsets using logistic and linear regression respectively. Consequence categories are stratified by rarity: moving from left to right the putatively damaging nature of the variants reduces from dark red to pink according to the legend, and the rarity reduces from a variant with MAC ≤ 5 in a *P*LI ≥ 0.9 gene and not in the non-psych portion of gnomAD (Not in gnomAD *p*LI ≥ 0.9), to a variant with MAC ≤ 5 (All) according to the *x*-axis labelling. Bars in panels B and C represent the 95% confidence intervals on the logistic and linear regression estimate of the enrichment of the class of variation labelled on the *x*-axis respectively. Regressions are run as described in supplementary materials: exome-wide burden analyses, and include sex, 10 PCs and total MAC ≤ 5 coding burden as covariates. Nominally significant enrichments or excess variants in cases are labelled with the associated *P-*value.

Given the excess burden observed between BD case status and ultra-rare PTV burden, we looked to tease apart this signal. We evaluated age at first impairment for a subset of 3,134 cases (supplementary materials: age of onset definitions, Table S6), but found no difference in the distribution of PTV burden or carrier status between earlier onset cases compared to older onset cases (minimum *P-*value across 50 tests using Kolmogorov-Smirnov (KS) test was 0.40, minimum *P-*value across 50 tests using Fisher’s exact tests 0.067 (supplementary materials: testing for relationship between age of onset and rare variant burden).

We also assessed whether the presence or absence of psychosis in a subset of 8,017 case samples (4,214 with psychosis (comprising 3,152 BD1, 661 BD2, 352 BDNOS, and 49 BD without a fine subclassification), 3,803 without psychosis (comprising 1,423 BD1, 1,845 BD2, 505 BDNOS, and 30 BD without a fine subclassification)) stratified risk (Table S7, supplementary materials: psychosis definitions). Both case subsets displayed significant enrichment of ultra-rare PTV burden in constrained genes (OR = 1.12, *P =* 0.0018; OR = 1.16, *P =* 6.6 × 10^−5^ for cases with and without psychosis respectively). However, there was no significant difference in excess burden between the two subcategories when including psychosis status as a covariate in the regression of ultra-rare PTV burden in constrained genes on case status (*P =* 0.42).

Restricting to missense enrichment, we do not observe a significant signal of enrichment of highly putatively damaging missense (MPC > 2) variation in bipolar disorder cases, in contrast to schizophrenia (*19*); Figure 1B,C. However, outside constrained genes, we observe significant enrichment of ultra-rare damaging missense variation across both BD subtypes with BD2 showing the strongest enrichment; Figure 1B,C (BD: OR = 1.02, *P =* 0.014; BD1: OR = 1.02, *P* = 0.014; BD2: OR = 1.03, *P* = 0.0036).

### Candidate gene-set and tissue enrichment

Beyond exome-wide and constrained gene burden, biologically and empirically informed gene sets can refine our understanding of how ultra-rare PTVs confer risk for BD and generate potential biological hypotheses for follow-up analyses. Using the Genotype-Tissue Expression portal (*20*), we find weak evidence for enrichment of ultra-rare PTVs in 13,372 genes expressed in brain tissues in bipolar cases; OR = 1.01, *P =* 0.032, compared to genes expressed in non-brain tissues; 23,450 genes, OR = 1.00, *P =* 0.15. More broadly, we tested for enrichment of ultra-rare PTVs in 43 GTEx tissues ((*21*), Table S8) defined as having the strongest tissue specific expression (Figure 2A, Figure S6). The pattern of enrichment for damaging ultra-rare variation resides predominantly in brain tissues, with the strongest association seen in the Amygdala (OR = 1.03, *P =* 3.9 × 10^−5^), a brain region previously found to be reduced in size in BD1 cases (*22*).

**Figure 2:**
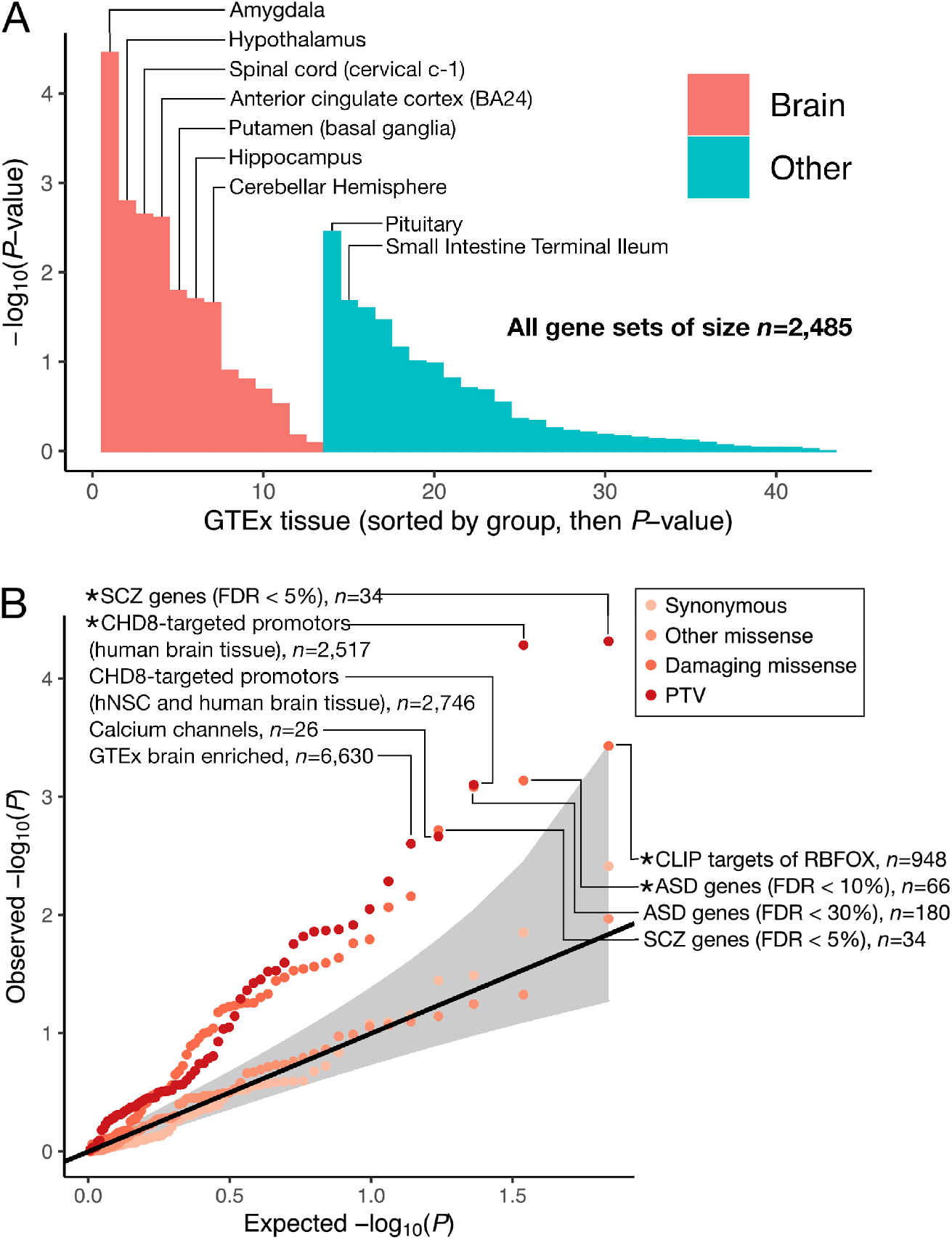
Biological insights from bipolar case-control whole-exome sequencing data. A. Enrichment of ultra-rare PTVs in BD cases over controls in tissue-specific expression genesets. Gene-sets are defined in (*21*) in detail. Bars are ordered first by whether they are a brain-tissue, and then by *P-*value. B. Enrichment of ultra-rare variants in targeted 68 gene-sets taken from the literature (*19, 29*). Top PTV and damaging missense gene-sets are labelled, and annotated with the number of genes in each geneset. Classes of variants tested in each gene-set are coloured according to the legend. Gene sets surpassing Bonferroni test correction are labelled with an asterisk.

We then considered 68 candidate gene-sets either generated or implicated in previous genetic studies of psychiatric disorders (supplementary materials: gene-set enrichment analysis, Figure 2B, Figure S7), and a more strictly defined collection of genes brain-enriched in GTEx: average expression over two-fold higher in brain tissues than the average across all tissues in GTEx (*23*). With this more stringent brain-enrichment definition (6,630 genes), we saw stronger ultra-rare PTV enrichment; OR = 1.04, *P* = 2.49 × 10^−3^. Among the 68 candidate gene sets, we observe significant enrichment (multiple test correction set at *P* < 7.35 × 10^−4^) of ultra-rare variation in four gene sets. For ultra-rare PTVs, we see significant enrichment in SCHEMA genes; FDR < 5% (*19*); 34 genes, OR = 1.89, *P =* 4.81 × 10^−5^, and CHD8 binding targets in human brain (*24*); 2,517 genes, OR = 1.09, *P =* 5.18 × 10^−5^. For ultra-rare damaging missense variants, we see significant enrichment in genes targeted by RBFOX (*25*); 948 genes, OR = 1.07, *P =* 3.70 × 10^−4^, and ASD FDR < 10% (*26*); 66 genes, OR = 1.24, *P =* 7.25 × 10^−4^. The enrichment of ultra-rare PTVs in SCHEMA and damaging missense in ASD provides further evidence of convergence of shared signal in the ultra-rare end of the allele frequency spectrum, mirroring the overlapping genetic risk for schizophrenia and BD observed in common variation (*27*), and schizophrenia and ASD in rare variation (*19*). Notably, we did not observe a rare-variant enrichment of damaging variation in gene sets generated from GWAS of BD (*3*). To investigate the rare-variant signal with schizophrenia further, we considered four distinct gene-sets of size 50, ordered by *P*-value in SCHEMA (*19*). Further PTV enrichment was observed in the top 50 genes over the FDR < 5% set; OR = 2.02, *P =* 8.14 × 10^−7^, but this significant enrichment was not observed as we moved down through the genes displaying ultra-rare damaging case-control enrichment in SCHEMA (genes 51-100; OR = 0.936, *P* = 0.680, genes 101-150; OR = 1.03, *P* = 0.794, genes 151-200; OR = 1.07, *P* = 0.686). We also did not observe PTV enrichment of ultra-rare damaging variation in the recently fine-mapped schizophrenia genes published by the PGC (*11*): OR = 1.10, *P* = 0.178.

Along with a candidate gene-set enrichment analysis approach, we considered a broad-based enrichment analysis using gene-sets derived from large pathway databases including Gene Ontology (GO), REACTOME and KEGG); a total of 1,697 gene-sets (Figure S8). By analysing excess rare variant burden in such a large collection of gene-lists we sought to elucidate pathways enriched for damaging variation associated with bipolar disorder in an agnostic manner. We observed significant enrichment of one gene-set after correction for multiple tests: genes involved in the G1/S transition of mitotic cell cycle; OR = 1.46, *P =* 1.36 × 10^−5^.

### Gene based-analysis approach

To boost power for gene discovery, we again restricted to ultra-rare variants which were not present in the non-psych portion of gnomAD (*28*), and we further enriched for pathogenic variants by restricting our analysis to ultra-rare variants (not in gnomAD non-psych, MAC ≤ 5) that are also either PTVs (Table S5) or damaging missense variants (supplementary materials: gene-based analysis approach; Table S5). Throughout, we use Fisher’s exact tests in each gene to test for case-control enrichment (supplementary materials: gene-based analysis approach, Figures S9-13).

### *AKAP11* implicated by ultra rare protein truncating variants

In our primary analysis, no gene surpassed genome-wide significance (set at *P* < 2.14 × 10^−6^ for 23,321 tests; dotted line in Figure 3), with the strongest case-control enrichment observed in *AKAP11* (*P* = 1.15 × 10^−5^ in BD, *P* = 5.30 × 10^−6^ in BD1). We do, however, begin to observe deviation from the null in the collection of tests of ultra rare PTV enrichment in bipolar cases, particularly in BD1 (Figure S14). This deviation was not observed for BD2 (Figure S15) despite the genome-wide enrichment of the PTV signal (Figure 1B,C), and is likely due to the reduced power of Fisher’s exact tests in BD2 case counts (*n* = 3,446).

**Figure 3:**
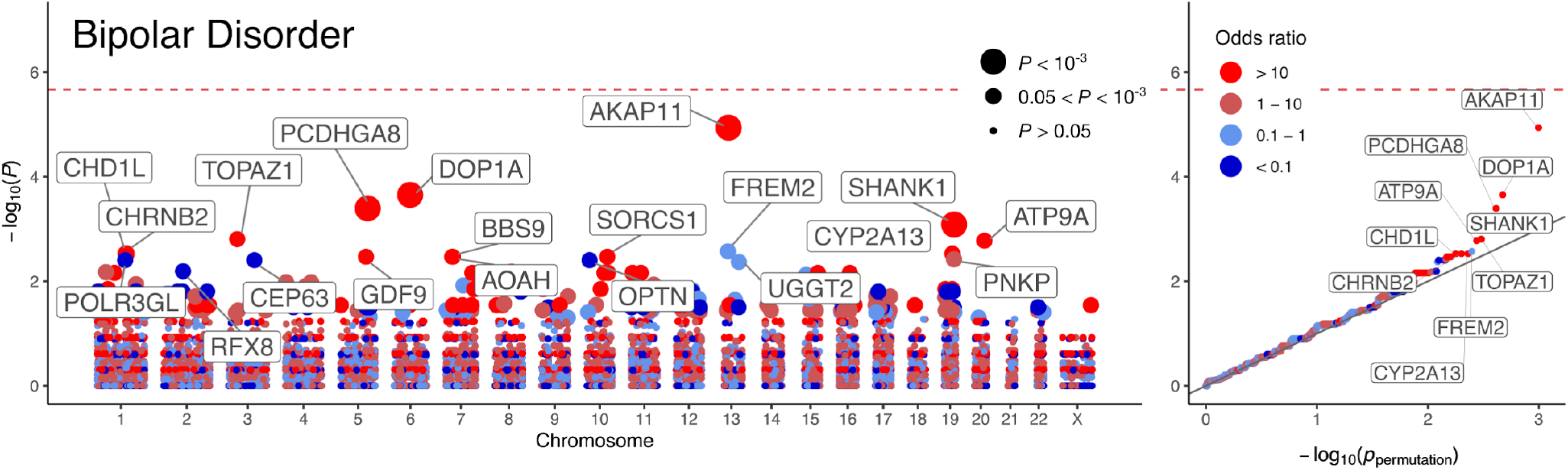
Results of the analysis of ultra-rare PTVs in 13,933 cases and 14,422 controls. Gene-based Manhattan and QQ plot for bipolar disorder (comprising BD1, BD2 and BDNOS). −log_10_ *P-*values obtained via Fisher’s exact tests are plotted against genetic position for each of the analysed genes. In the QQ plots, observed −log_10_ *P-*values are plotted against permutation *P-*values according to the procedure described in the supplementary materials: gene-based analysis approach. Points are coloured according to the discrete scale displayed in the legend. In the Manhattan plot and QQ plot, the gene symbols of the top 20 and top 10 genes by *P-*value are labelled, respectively. Points in the Manhattan plot are sized according to *P*-value as displayed in the legend.

Given the strong overlap in common variant risk between BD and schizophrenia, we sought to determine whether there is evidence of a shared signal of enrichment of ultra-rare PTVs in BD and schizophrenia cases. Due to overlap in controls between SCHEMA and BipEx, we analysed an ultra-rare variant count data-set which excluded these controls, and meta-analysed the data (supplementary materials: combining SCHEMA and BipEx data in meta-analysis). To avoid the schizophrenia ultra-rare PTV case-control enrichment signal overwhelming the BD signal when presenting results, we first sort on *P-*value in the primary gene-based BD analysis and display the top 10 *P-*values before and after meta-analysis with SCHEMA counts, Table 1 and Table S9. The combined analysis in BD and schizophrenia cases reveal one exome-wide significant gene, *AKAP11* (*P =* 2.83 × 10^−9^), and one suggestive gene, *ATP9A* (*P =* 5.36 × 10^−6^).

**Table 1:**
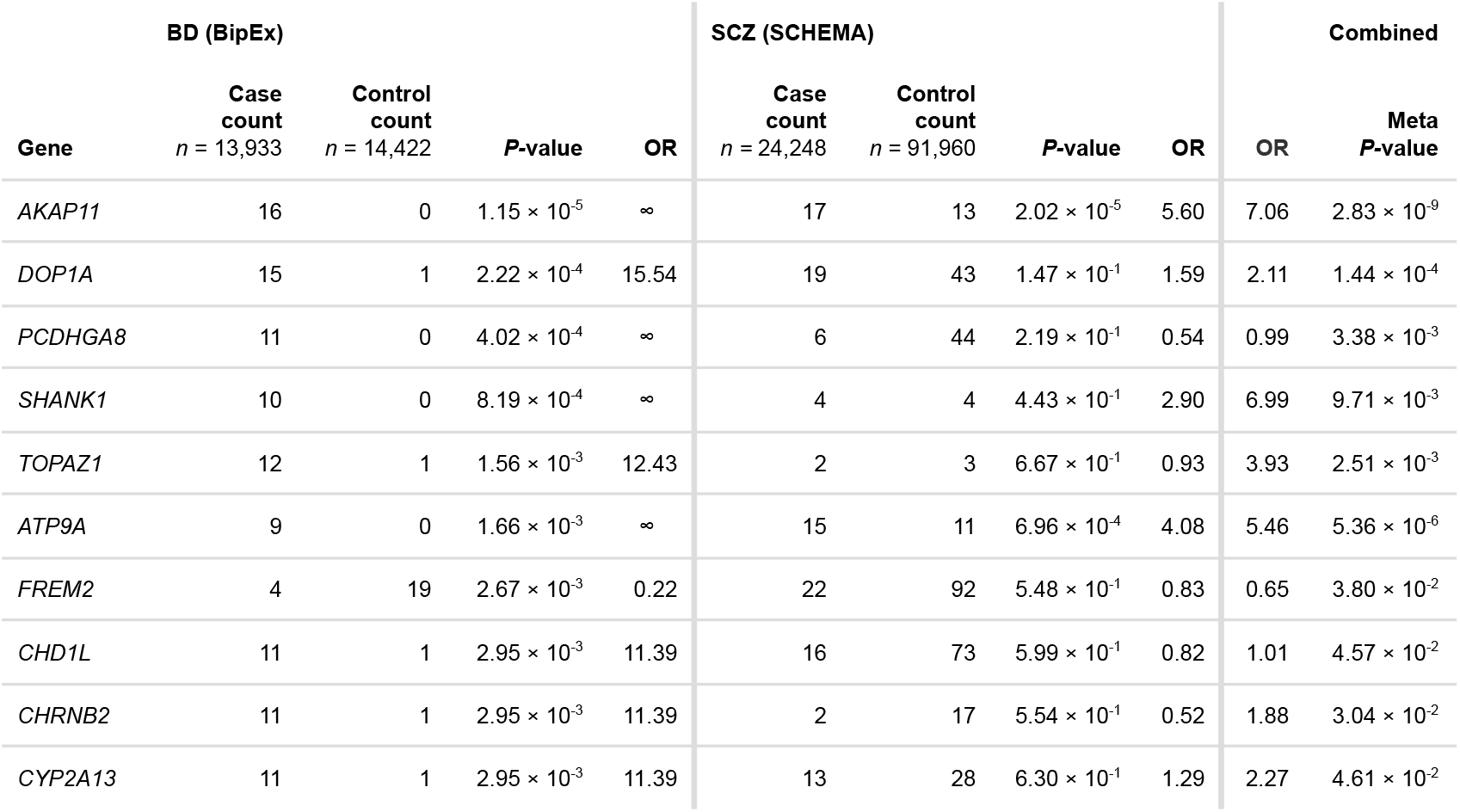
BipEx and SCHEMA case-control counts of the top ten most significant genes in the BipEx gene-based analysis. Case and control columns denote the count of ultra-rare PTVs in the gene in the respective dataset. *P-*values are determined using Fisher’s exact and CMH tests for BipEx and SCHEMA (supplementary materials: gene-based analysis approach) respectively, and meta-analysed weighting by effective sample size. BipEx: BD case count 13,933, control count 14,422. SCHEMA: schizophrenia case count 24,248, control count 91,960. The SCHEMA OR is the estimated OR averaged over strata, whereas the combined OR is the simple OR calculated by combining the BipEx and SCHEMA cases and controls. Note that differential coverage across exome sequencing platforms and whole genome sequencing means that case/control counts differ across genes.

The top gene hit, *AKAP11* (the gene encoding A-Kinase Anchoring Protein 11, also known as AKAP220) contains only a single isoform, is under evolutionary constraint (LOEUF = 0.3, *p*LI = 0.98), and is highly expressed in the brain (cerebellar hemisphere (38.54 median TPM), frontal cortex (BA9) (31.52 median TPM); (*20*)) and has been shown to interact with *GSK3B*, the hypothesized target of lithium therapy (*30*–*32*). Therefore, we gathered all available lithium response information for carriers of *AKAP11* PTVs among the BD cases (supplementary materials: lithium response). Of the eleven cases for which lithium response information was available, seven reported a good response (of which five were in SWEBIC cohort and reported ‘complete response, recovered’, and two were in the Cardiff collection and reported that lithium helped stabilise their moods), and four did not respond well to lithium. Of the poor responders, three were in the London cohort, and one was in the SWEBIC cohort. While the percent of good responders in *AKAP11* PTV carriers (63.6%) is marginally elevated relative to the background response rate in available BD cases (52%), the sample size is far too small to form any robust conclusions from the data.

The rare damaging association appears to be specific to BD and schizophrenia. To our knowledge, there is no signal of enrichment in *AKAP11* in other neurodevelopmental disorders. *AKAP11* is not present in the collection of ‘developmental disorder genes’ curated to be associated with developmental disorders (*33*); https://decipher.sanger.ac.uk/ddd/ddgenes), the autism sequencing consortium (ASC) analysis (*34*), or Epi25 study (*35*). Furthermore, expression of *AKAP11* tends to occur later in development (Figure S16).

We also examined ultra-rare PTV variant counts in the Bipolar Sequencing Consortium (BSC) (*14*) (supplementary materials: external validation with the BSC exome data, Table S10) exome sequence data. Non-zero count data were available for seven of the top ten genes exhibiting differences in ultra-rare PTV counts between BD cases and controls as measured by *P-*value in the BipEx dataset. Of these, one was enriched for ultra-rare PTVs in controls (FREM2) in BipEx, and did not display control enrichment in the BSC data. The remaining six displayed case-control enrichment in BipEx. In four out of these six genes (including *AKAP11* and *ATP9A*), we observed further case enrichment (Table S11) in the BSC data.

## Discussion

In the largest bipolar disorder exome study to date, ultra rare PTVs in constrained genes are significantly enriched in BD cases. In fact, enrichment in constrained genes remains significant even after excluding the top 20 BD-risk associated genes (OR = 1.07; *P =* 0.00313) with *p*LI ≥ 0.9 (Table S12) reflecting the highly polygenic genetic architecture of bipolar disorder, a property shared with schizophrenia (*19*), and suggesting that the majority of genes involved in bipolar disorder risk will require larger sample sizes to be discovered. Furthermore, ultra rare PTVs are significantly enriched in schizophrenia risk genes identified in the SCHEMA consortium, suggesting that rare variation in these genes are not distinct to schizophrenia pathophysiology, and that overlap in risk for schizophrenia and BD is now evident in both rare and common variation. Finally, combining our results with data from SCHEMA reveals strong evidence that haploinsufficiency in *AKAP11* confers risk for both BD and schizophrenia, but not for early-onset neurodevelopmental disorders.

*AKAP11* codes for the AKAP-11 protein (also known as AKAP220), one of a family of scaffolding proteins that bind to the regulatory subunit of the protein kinase A (PKA). These anchoring proteins confine PKA to discrete locations in the cell to target specific substrates for phosphorylation and dephosphorylation. In particular, GSK3B is bound by AKAP-11. GSK3B is hypothesized to be the target of lithium, the primary treatment for bipolar disorder (*36*). By binding to GSK3B, AKAP-11 mediates PKA-dependent inhibition of GSK3B - PKA inhibits the activity of GSK3B bound to AKAP-11 more strongly than GSK3B in general, and thus modifications to AKAP-11 have the potential to affect downstream pathways. *GSK3B* is one of two paralogous genes (*GSK3* alpha and *GSK3* beta) that encode a serine/threonine protein kinase, Glycogen synthase kinase 3. The primary known function of which is phosphorylation of more than one hundred different substrates, thus affecting a myriad of signalling pathways (*4, 36, 37*). With this in mind, we looked to determine the efficacy of lithium treatment in BD cases harboring an ultra-rare PTV in *AKAP11*. Of the eleven individuals with treatment data available, seven displayed a good response to lithium treatment, in line with the notion that the effects of disrupting AKAP-11 may be partially rescued by lithium therapy. However, the ultra-rare PTV carrier sample size is currently too low to draw robust conclusions regarding lithium treatment response.

Beyond PTV enrichment in constrained genes, we see early evidence of enrichment in ultra-rare damaging missense variation, particularly within BD2. This enrichment is evident outside of missense constrained regions (as defined by MPC > 2), which is perhaps surprising given the signal of association seen in this slice of rare variation in schizophrenia cases is mainly within constrained missense regions. Because BD2 displays a stronger correlation of common variant effects with major depression, with BD1 more correlated to schizophrenia, there is a chance that this missense signal is capturing something distinct to mood disorders relative to psychotic disorders. However, we should be cautious not to read too much into differences in ultra-rare damaging missense enrichment across the BD subtypes; the number of BD2 samples (*n* = 3,446) in the BipEx dataset is less than half of BD1 (*n* = 8,238), and confidence intervals around the damaging missense enrichment overlap (Figure 1). Furthermore, attempts to refine this exome-wide signal to individual genes or targeted gene sets did not result in any significant signals of association after correcting for multiple testing (Figure S15, Figure S7). As with PTV enrichment, we expect to see a refinement of the putatively damaging missense signal as sample sizes increase.

Despite sequencing 13,933 BD cases, we did not observe any BD specific risk genes surpassing exome-wide significance. In contrast, the 24,248 schizophrenia cases analyzed in SCHEMA have yielded 10 significant risk genes. When we compare the observed ultra-rare PTV enrichment among constrained genes in our current sample (OR = 1.11) to SCHEMA (OR = 1.26), we currently estimate that about double the case sample size of schizophrenia is needed in BD to achieve comparable statistical power to discover individual risk genes. Moreover, schizophrenia has now seen meaningful convergence of gene overlap in the common and rare end of the allele frequency spectrum, in large part through larger exome sample sizes as well as fine-mapping of GWAS loci (*19*). The overlap in BD, however, remains uncertain. The BSC exome-dataset examined 3,987 BD cases (*14*), finding suggestive enrichment in 165 genes implicated in BD GWAS (OR = 1.9, *P =* 6.0 × 10^−4^), but this finding did not replicate in the current sample (OR = 0.9, *P =* 0.40). Given that prior evidence of common and rare gene overlap in schizophrenia was quite modest (*16, 17, 29*), as sample sizes increase for both common and rare variation analyses in BD, we do expect to see a slow but steady convergence in much the same manner.

In summary, ultra-rare PTVs in constrained genes are significantly enriched in BD patients over controls, a result firmly established in schizophrenia and other early-onset neurodevelopmental disorders. We are beginning to see promising signals among individual genes, despite not having any surpassing exome-wide significance for BD alone. We observe that shared risk for bipolar disorder and schizophrenia is present in both common and damaging ultra-rare variation. Our top gene, *AKAP11*, shows shared evidence of risk for BD and schizophrenia, increasing our confidence that we are discovering true risk factors underlying psychiatric disease. Overall, the current evidence suggests BD is on a similar trajectory to schizophrenia, where increasing sample sizes and further collaborative efforts will inevitably lead to biologically meaningful risk genes and pathways underlying BD risk.

## Data Availability

We have provided data tables in the main text, Supplementary Methods. and in an exome results browser at https://bipex.broadinstitute.org. We display all of our results, from the variant and gene level in a browser available at https://bipex.broadinstitute.org. A detailed summary of phenotype curation, and QC, including additional plots is available at https://astheeggeggs.github.io/BipEx/. Whole Exome Sequence data generated under this study are hosted on and shared with the collaborating study groups via the controlled access Terra platform (https://app.terra.bio). The Terra environment, created by the Broad Institute, contains a rich system of workspace functionalities centered on data sharing and analysis. Requests for access to the controlled datasets are managed by data custodians at the Broad Institute and sent to sample contributing investigators for approval.

## Data availability

We display all of our results, from the variant and gene level in a browser available at https://bipex.broadinstitute.org. A detailed summary of phenotype curation, and QC, including additional plots is available at https://astheeggeggs.github.io/BipEx/. Whole Exome Sequence data generated under this study are hosted on and shared with the collaborating study groups via the controlled access Terra platform (https://app.terra.bio). The Terra environment, created by the Broad Institute, contains a rich system of workspace functionalities centered on data sharing and analysis. Requests for access to the controlled datasets are managed by data custodians at the Broad Institute and sent to sample contributing investigators for approval.

## Ethics statement

IRB approvals and study consent forms from each of sample contributing organizations were sent to the Broad Institute before samples were sequenced and analyzed. Contributing organizations include: University of Aberdeen, Trinity College Dublin, University of Edinburgh, University College London, Cardiff University, University of Cambridge, Vrije Universitat Amsterdam, University College of Los Angeles, Universitats Klinikum Frankfurt, Massachusetts General Hospital, Johns Hopkins University, Karolinska Institute, LifeGene Biorepository at Karolinska Institute, and Umea University.

All ethical approvals are on file at the Massachusetts General Brigham (MGB), formerly Partners, IRB office amended to protocol #2014P001342, title: ‘Molecular Profiling of Psychiatric Disease’.

## Code availability

Code used to perform QC, analysis, and creation of plots is provided at github.com/astheeggeggs/BipEx.

## Acknowledgements

This study was supported by generous support from the Stanley Family Foundation, Kent and Elizabeth Dauten, and The Dalio Foundation who have enabled us to rapidly expand our data generation collections with the goal of moving towards better treatments for bipolar disorder, schizophrenia, and other psychiatric disorders. BSC grant support was provided by the NIH, grant R01MH110437. We thank Willem Ouwehand for contributing control samples for exome sequencing.

## Competing interests

B.M.N. is a member of the scientific advisory board at Deep Genomics and consultant for Camp4 Therapeutics, Takeda Pharmaceutical, and Biogen. D.S.P. is an employee of Genomics plc. All the analyses reported in this paper were performed as part of D.S.P.’s previous employment at the Analytic and Translational Genetics Unit, Department of Medicine, Massachusetts General Hospital, Boston, Massachusetts, USA, and Stanley Center for Psychiatric Research, Broad Institute of MIT and Harvard, Cambridge, Massachusetts, USA.. C-Y.C. is an employee of Biogen. F.D. is an employee of Sheppard Pratt. A.L. and E.A.S. are now employees of Regeneron. All other authors declare no competing interests.

## Notes

### Author Declarations

IRB approvals and study consent forms from each of sample contributing organizations were sent to the Broad Institute before samples were sequenced and analyzed. Contributing organizations include: University of Aberdeen, Trinity college Dublin, University of Edinburgh, University College London, Cardiff University, University of Cambridge, Vrije Universitat, Amsterdam, University College of Los Angeles, Universitats Klinikum Frankfurt, Massachusetts General Hospital, Johns Hopkins University, Karolinska Institute, LifeGene Biorepository at Karolinska Institute, and Umea University. All ethical approvals are on file at the Massachusetts General Brigham (MGB), formerly Partners, IRB office amended to protocol #2014P001342, title: 'Molecular Profiling of Psychiatric Disease'.

### Summary of Updates

Revised author list

## References

1. A. J. Ferrari, E. Stockings, J.-P. Khoo, H. E. Erskine, L. Degenhardt, T. Vos, H. A. Whiteford, The prevalence and burden of bipolar disorder: findings from the Global Burden of Disease Study 2013. Bipolar Disord. 18, 440–450 (2016).

2. T. J. C. Polderman, B. Benyamin, C. A. de Leeuw, P. F. Sullivan, A. van Bochoven, P. M. Visscher, D. Posthuma, Meta-analysis of the heritability of human traits based on fifty years of twin studies. Nat. Genet. 47, 702–709 (2015).

3. E. A. Stahl, G. Breen, A. J. Forstner, A. McQuillin, S. Ripke, V. Trubetskoy, M. Mattheisen, Y. Wang, J. R. I. Coleman, H. A. Gaspar, C. A. de Leeuw, S. Steinberg, J. M. W. Pavlides, M. Trzaskowski, E. M. Byrne, T. H. Pers, P. A. Holmans, A. L. Richards, L. Abbott, E. Agerbo, H. Akil, D. Albani, N. Alliey-Rodriguez, T. D. Als, A. Anjorin, V. Antilla, S. Awasthi, J. A. Badner, M. Bækvad-Hansen, J. D. Barchas, N. Bass, M. Bauer, R. Belliveau, S. E. Bergen, C. B. Pedersen, E. Bøen, M. P. Boks, J. Boocock, M. Budde, W. Bunney, M. Burmeister, J. Bybjerg-Grauholm, W. Byerley, M. Casas, F. Cerrato, P. Cervantes, K. Chambert, A. W. Charney, D. Chen, C. Churchhouse, T.-K. Clarke, W. Coryell, D. W. Craig, C. Cruceanu, D. Curtis, P. M. Czerski, A. M. Dale, S. de Jong, F. Degenhardt, J. Del-Favero, J. R. DePaulo, S. Djurovic, A. L. Dobbyn, A. Dumont, T. Elvsåshagen, V. Escott-Price, C. C. Fan, S. B. Fischer, M. Flickinger, T. M. Foroud, L. Forty, J. Frank, C. Fraser, N. B. Freimer, L. Frisén, K. Gade, D. Gage, J. Garnham, C. Giambartolomei, M. G. Pedersen, J. Goldstein, S. D. Gordon, K. Gordon-Smith, E. K. Green, M. J. Green, T. A. Greenwood, J. Grove, W. Guan, J. Guzman-Parra, M. L. Hamshere, M. Hautzinger, U. Heilbronner, S. Herms, M. Hipolito, P. Hoffmann, D. Holland, L. Huckins, S. Jamain, J. S. Johnson, A. Juréus, R. Kandaswamy, R. Karlsson, J. L. Kennedy, S. Kittel-Schneider, J. A. Knowles, M. Kogevinas, A. C. Koller, R. Kupka, C. Lavebratt, J. Lawrence, W. B. Lawson, M. Leber, P. H. Lee, S. E. Levy, J. Z. Li, C. Liu, S. Lucae, A. Maaser, D. J. MacIntyre, P. B. Mahon, W. Maier, L. Martinsson, S. McCarroll, P. McGuffin, M. G. McInnis, J. D. McKay, H. Medeiros, S. E. Medland, F. Meng, L. Milani, G. W. Montgomery, D. W. Morris, T. W. Mühleisen, N. Mullins, H. Nguyen, C. M. Nievergelt, A. N. Adolfsson, E. A. Nwulia, C. O’Donovan, L. M. O. Loohuis, A. P. S. Ori, L. Oruc, U. Ösby, R. H. Perlis, A. Perry, A. Pfennig, J. B. Potash, S. M. Purcell, E. J. Regeer, A. Reif, C. S. Reinbold, J. P. Rice, F. Rivas, M. Rivera, P. Roussos, D. M. Ruderfer, E. Ryu, C. Sánchez-Mora, A. F. Schatzberg, W. A. Scheftner, N. J. Schork, C. Shannon Weickert, T. Shehktman, P. D. Shilling, E. Sigurdsson, C. Slaney, O. B. Smeland, J. L. Sobell, C. Søholm Hansen, A. T. Spijker, D. St Clair, M. Steffens, J. S. Strauss, F. Streit, J. Strohmaier, S. Szelinger, R. C. Thompson, T. E. Thorgeirsson, J. Treutlein, H. Vedder, W. Wang, S. J. Watson, T. W. Weickert, S. H. Witt, S. Xi, W. Xu, A. H. Young, P. Zandi, P. Zhang, S. Zöllner, eQTLGen Consortium, BIOS Consortium, R. Adolfsson, I. Agartz, M. Alda, L. Backlund, B. T. Baune, F. Bellivier, W. H. Berrettini, J. M. Biernacka, D. H. R. Blackwood, M. Boehnke, A. D. Børglum, A. Corvin, N. Craddock, M. J. Daly, U. Dannlowski, T. Esko, B. Etain, M. Frye, J. M. Fullerton, E. S. Gershon, M. Gill, F. Goes, M. Grigoroiu-Serbanescu, J. Hauser, D. M. Hougaard, C. M. Hultman, I. Jones, L. A. Jones, R. S. Kahn, G. Kirov, M. Landén, M. Leboyer, C. M. Lewis, Q. S. Li, J. Lissowska, N. G. Martin, F. Mayoral, S. L. McElroy, A. M. McIntosh, F. J. McMahon, I. Melle, A. Metspalu, P. B. Mitchell, G. Morken, O. Mors, P. B. Mortensen, B. Müller-Myhsok, R. M. Myers, B. M. Neale, V. Nimgaonkar, M. Nordentoft, M. M. Nöthen, M. C. O’Donovan, K. J. Oedegaard, M. J. Owen, S. A. Paciga, C. Pato, M. T. Pato, D. Posthuma, J. A. Ramos-Quiroga, M. Ribasés, M. Rietschel, G. A. Rouleau, M. Schalling, P. R. Schofield, T. G. Schulze, A. Serretti, J. W. Smoller, H. Stefansson, K. Stefansson, E. Stordal, P. F. Sullivan, G. Turecki, A. E. Vaaler, E. Vieta, J. B. Vincent, T. Werge, J. I. Nurnberger, N. R. Wray, A. Di Florio, H. J. Edenberg, S. Cichon, R. A. Ophoff, L. J. Scott, O. A. Andreassen, J. Kelsoe, P. Sklar, Bipolar Disorder Working Group of the Psychiatric Genomics Consortium, Genome-wide association study identifies 30 loci associated with bipolar disorder. Nat. Genet. 51, 793–803 (2019).

4. A. W. Charney, E. A. Stahl, E. K. Green, C.-Y. Chen, J. L. Moran, K. Chambert, R. A. Belliveau Jr, L. Forty, K. Gordon-Smith, P. H. Lee, E. J. Bromet, P. F. Buckley, M. A. Escamilla, A. H. Fanous, L. J. Fochtmann, D. S. Lehrer, D. Malaspina, S. R. Marder, C. P. Morley, H. Nicolini, D. O. Perkins, J. J. Rakofsky, M. H. Rapaport, H. Medeiros, J. L. Sobell, L. Backlund, S. E. Bergen, A. Juréus, M. Schalling, P. Lichtenstein, J. A. Knowles, K. E. Burdick, I. Jones, L. A. Jones, C. M. Hultman, R. Perlis, S. M. Purcell, S. A. McCarroll, C. N. Pato, M. T. Pato, A. Di Florio, N. Craddock, M. Landén, J. W. Smoller, D. M. Ruderfer, P. Sklar, Contribution of Rare Copy Number Variants to Bipolar Disorder Risk Is Limited to Schizoaffective Cases. Biol. Psychiatry. 86, 110–119 (2019).

5. A. Ganna, F. K. Satterstrom, S. M. Zekavat, I. Das, M. I. Kurki, C. Churchhouse, J. Alfoldi, A. R. Martin, A. S. Havulinna, A. Byrnes, W. K. Thompson, P. R. Nielsen, K. J. Karczewski, E. Saarentaus, M. A. Rivas, N. Gupta, O. Pietiläinen, C. A. Emdin, F. Lescai, J. Bybjerg-Grauholm, J. Flannick, GoT2D/T2D-GENES Consortium, J. M. Mercader, M. Udler, SIGMA Consortium Helmsley IBD Exome Sequencing Project, FinMetSeq Consortium, iPSYCH-Broad Consortium, M. Laakso, V. Salomaa, C. Hultman, S. Ripatti, E. Hämäläinen, J. S. Moilanen, J. Körkkö, O. Kuismin, M. Nordentoft, D. M. Hougaard, O. Mors, T. Werge, P. B. Mortensen, D. MacArthur, M. J. Daly, P. F. Sullivan, A. E. Locke, A. Palotie, A. D. Børglum, S. Kathiresan, B. M. Neale, Quantifying the Impact of Rare and Ultra-rare Coding Variation across the Phenotypic Spectrum. Am. J. Hum. Genet. 102, 1204–1211 (2018).

6. A. J. J. F. Crow, M. Kimura, An Introduction to Population Genetics Theory. Population (French Edition). 26 (1971), p. 977.

7. G. V. Kryukov, L. A. Pennacchio, S. R. Sunyaev, Most rare missense alleles are deleterious in humans: implications for complex disease and association studies. Am. J. Hum. Genet. 80, 727–739 (2007).

8. R. A. Power, S. Kyaga, R. Uher, J. H. MacCabe, N. Långström, M. Landen, P. McGuffin, C. M. Lewis, P. Lichtenstein, A. C. Svensson, Fecundity of patients with schizophrenia, autism, bipolar disorder, depression, anorexia nervosa, or substance abuse vs their unaffected siblings. JAMA Psychiatry. 70, 22–30 (2013).

9. American Psychiatric Association. Task Force on DSM-IV., DSM-IV Sourcebook (Amer Psychiatric Pub Incorporated, 1998).

10. A. Janca, T. B. Ustün, T. S. Early, N. Sartorius, The ICD-10 symptom checklist: a companion to the ICD-10 classification of mental and behavioural disorders. Soc. Psychiatry Psychiatr. Epidemiol. 28, 239–242 (1993).

11. N. Mullins, A. J. Forstner, K. S. O’Connell, B. Coombes, J. R. I. Coleman, Z. Qiao, T. D. Als, T. B. Bigdeli, S. Børte, J. Bryois, A. W. Charney, O. K. Drange, M. J. Gandal, S. P. Hagenaars, M. Ikeda, N. Kamitaki, M. Kim, K. Krebs, G. Panagiotaropoulou, B. M. Schilder, L. G. Sloofman, S. Steinberg, V. Trubetskoy, B. S. Winsvold, H.-H. Won, L. Abramova, K. Adorjan, E. Agerbo, M. Al Eissa, D. Albani, N. Alliey-Rodriguez, A. Anjorin, V. Antilla, A. Antoniou, S. Awasthi, J. Hyun Baek, M. Bækvad-Hansen, N. Bass, M. Bauer, E. C. Beins, S. E. Bergen, A. Birner C. Bøcker Pedersen, E. Bøen, M. P. Boks, R. Bosch, M. Brum, B. M. Brumpton, N. Brunkhorst-Kanaan, M. Budde, J. Bybjerg-Grauholm, W. Byerley, M. Cairns, M. Casas, P. Cervantes, T.-K. Clarke, C. Cruceanu, A. Cuellar-Barboza, J. Cunningham, D. Curtis, P. M. Czerski, A. M. Dale, N. Dalkner, F. S. David, F. Degenhardt, S. Djurovic, A. L. Dobbyn, A. Douzenis, T. Elvsåshagen, V. Escott-Price, I. Nicol Ferrier, A. Fiorentino, T. M. Foroud, L. Forty, J. Frank, O. Frei, N. B. Freimer, L. Frisén, K. Gade, J. Garnham, J. Gelernter, M. Marianne Giørtz Pedersen, I. R. Gizer, S. D. Gordon, K. Gordon-Smith, T. A. Greenwood, J. Grove, J. Guzman-Parra, K. Ha, M. Haraldsson, M. Hautzinger, U. Heilbronner, D. Hellgren, S. Herms, P. Hoffmann, P. A. Holmans, L. Huckins, S. Jamain, J. S. Johnson, J. L. Kalman, Y. Kamatani, J. L. Kennedy, S. Kittel-Schneider, J. A. Knowles, M. Kogevinas, M. Koromina, T. M. Kranz, H. R. Kranzler, M. Kubo, R. Kupka, S. A. Kushner, C. Lavebratt, J. Lawrence, M. Leber, H.-J. Lee, P. H. Lee, S. E. Levy, C. Lewis, C. Liao, S. Lucae, M. Lundberg, D. J. MacIntyre, W. Maier, A. Maihofer, D. Malaspina, E. Maratou, L. Martinsson, M. Mattheisen, N. W. McGregor, P. McGuffin, J. D. McKay, H. Medeiros, S. E. Medland, V. Millischer, G. W. Montgomery, J. L. Moran, D. W. Morris, T. W. Mühleisen, N. O’Brien, C. O’Donovan, L. M. Olde Loohuis, L. Oruc, S. Papiol, A. F. Pardiñas, A. Perry, A. Pfennig, E. Porichi, J. B. Potash, D. Quested, T. Raj, M. H. Rapaport, J. R. DePaulo, E. J. Regeer, J. P. Rice, F. Rivas, M. Rivera, J. Roth, P. Roussos, D. M. Ruderfer, C. Sánchez-Mora, E. C. Schulte, F. Senner, S. Sharp, P. D. Shilling, E. Sigurdsson, L. Sirignano, C. Slaney, O. B. Smeland, J. L. Sobell, C. S. Hansen, M. S. Artigas, A. T. Spijker, D. J. Stein, J. S. Strauss, B. Beata Światkowska, C. Terao, T. E. Thorgeirsson, C. Toma, P. Tooney, E.-E. Tsermpini, M. P. Vawter, H. Vedder, J. T. R. Walters, S. H. Witt, S. Xi, W. Xu, H. Young, A. H. Young, P. P. Zandi, H. Zhou, L. Zillich, R. Adolfsson, I. Agartz, M. Alda, L. Alfredsson, G. Babadjanova, L. Backlund, B. T. Baune, F. Bellivier, S. Bengesser, W. H. Berrettini, D. H. R. Blackwood, M. Boehnke, A. D. Børglum, G. Breen, V. J. Carr, S. Catts, A. Corvin, N. Craddock, U. Dannlowski, D. Dikeos, T. Esko, B. Etain, P. Ferentinos, M. Frye, J. M. Fullerton, M. Gawlik, E. S. Gershon, F. Goes, M. J. Green, M. Grigoroiu-Serbanescu, J. Hauser, F. Henskens, J. Hillert, K. S. Hong, D. M. Hougaard, C. M. Hultman, K. Hveem, N. Iwata, A. V. Jablensky, I. Jones, L. A. Jones, R. S. Kahn, J. R. Kelsoe, G. Kirov, M. Mikael Landén, M. Leboyer, C. M. Lewis, Q. S. Li, J. Lissowska, C. Lochner, C. Loughland, N. G. Martin, C. A. Mathews, F. Mayoral, S. L. McElroy, A. M. McIntosh, F. J. McMahon, I. Melle, P. Michie, L. Milani, P. B. Mitchell, G. Morken, O. Mors, P. Bo Mortensen, B. Mowry, B. Müller-Myhsok, R. M. Myers, B. M. Neale, C. M. Nievergelt, M. Nordentoft, M. M. Nöthen, M. C. O’Donovan, K. J. Oedegaard, T. Olsson, M. J. Owen, S. A. Paciga, C. Pantelis, C. Pato, M. T. Pato, G. P. Patrinos, R. H. Perlis, D. Posthuma, J. A. Ramos-Quiroga, A. Reif, E. Z. Reininghaus, M. Ribasés, M. Rietschel, S. Ripke, G. A. Rouleau, T. Saito, U. Schall, M. Schalling, P. R. Schofield, T. G. Schulze, R. J. Scott, L. J. Scott, A. Serretti, C. S. Weickert, J. W. Smoller, H. Stefansson, K. Stefansson, E. Stordal, F. Streit, P. F. Sullivan, G. Turecki, A. E. Vaaler, E. Vieta, J. B. Vincent, I. D. Waldman, T. W. Weickert, T. Werge, N. R. Wray, J.-A. Zwart, J. M. Biernacka, J. I. Nurnberger, S. Cichon, H. J. Edenberg, E. A. Stahl, A. McQuillin, A. Di Florio, R. A. Ophoff, O. A. Andreassen, HUNT All-In Psychiatry, Genome-wide association study of over 40,000 bipolar disorder cases provides novel biological insights (2020),, doi:10.1101/2020.09.17.20187054.

12. T. Husson, J.-B. Duboc, O. Quenez, C. Charbonnier, M. Rotharmel, M. Cuenca, X. Jegouzo, A.-C. Richard, T. Frebourg, J.-F. Deleuze, A. Boland, E. Genin, S. Debette, C. Tzourio, D. Campion, G. Nicolas, O. Guillin, FREX Consortium, Identification of potential genetic risk factors for bipolar disorder by whole-exome sequencing. Transl. Psychiatry. 8, 268 (2018).

13. J. H. Sul, S. K. Service, A. Y. Huang, V. Ramensky, S.-G. Hwang, T. M. Teshiba, Y. Park, A. P. S. Ori, Z. Zhang, N. Mullins, L. M. Olde Loohuis, S. C. Fears, C. Araya, X. Araya, M. Spesny, J. Bejarano, M. Ramirez, G. Castrillón, J. Gomez-Makhinson, M. C. Lopez, G. Montoya, C. P. Montoya, I. Aldana, J. I. Escobar, J. Ospina-Duque, B. Kremeyer, G. Bedoya, A. Ruiz-Linares, R. M. Cantor, J. Molina, G. Coppola, R. A. Ophoff, G. Macaya, C. Lopez-Jaramillo, V. Reus, C. E. Bearden, C. Sabatti, N. B. Freimer, Contribution of common and rare variants to bipolar disorder susceptibility in extended pedigrees from population isolates. Transl. Psychiatry. 10, 74 (2020).

14. X. Jia, F. S. Goes, A. E. Locke, D. Palmer, W. Wang, S. Cohen-Woods, G. Genovese, A. U. Jackson, C. Jiang, M. Kvale, N. Mullins, H. Nguyen, M. Pirooznia, M. Rivera, D. M. Ruderfer, L. Shen, K. Thai, M. Zawistowski, Y. Zhuang, G. Abecasis, H. Akil, S. Bergen, M. Burmeister, S. Champion, M. DelaBastide, A. Juréus, H. M. Kang, P.-Y. Kwok, J. Z. Li, S. E. Levy, E. T. Monson, J. Moran, J. Sobell, S. Watson, V. Willour, S. Zöllner, R. Adolfsson, D. Blackwood, M. Boehnke, G. Breen, A. Corvin, N. Craddock, A. DiFlorio, C. M. Hultman, M. Landen, C. Lewis, S. A. McCarroll, W. Richard McCombie, P. McGuffin, A. McIntosh, A. McQuillin, D. Morris, R. M. Myers, M. O’Donovan, R. Ophoff, M. Boks, R. Kahn, W. Ouwehand, M. Owen, C. Pato, M. Pato, D. Posthuma, J. B. Potash, A. Reif, P. Sklar, J. Smoller, P. F. Sullivan, J. Vincent, J. Walters, B. Neale, S. Purcell, N. Risch, C. Schaefer, E. A. Stahl, P. P. Zandi, L. J. Scott, Investigating rare pathogenic/likely pathogenic exonic variation in bipolar disorder. Mol. Psychiatry (2021), doi:10.1038/s41380-020-01006-9.

15. C. Cruceanu, J.-F. Schmouth, S. G. Torres-Platas, J. P. Lopez, A. Ambalavanan, E. Darcq, F. Gross, B. Breton, D. Spiegelman, D. Rochefort, P. Hince, J. M. Petite, J. Gauthier, R. G. Lafrenière, P. A. Dion, C. M. Greenwood, B. L. Kieffer, M. Alda, G. Turecki, G. A. Rouleau, Rare susceptibility variants for bipolar disorder suggest a role for G protein-coupled receptors. Molecular Psychiatry. 23 (2018), pp. 2050–2056.

16. G. Genovese, M. Fromer, E. A. Stahl, D. M. Ruderfer, K. Chambert, M. Landén, J. L. Moran, S. M. Purcell, P. Sklar, P. F. Sullivan, C. M. Hultman, S. A. McCarroll, Increased burden of ultra-rare protein-altering variants among 4,877 individuals with schizophrenia. Nature Neuroscience. 19 (2016), pp. 1433–1441.

17. T. Singh, J. T. R. Walters, M. Johnstone, D. Curtis, J. Suvisaari, M. Torniainen, E. Rees, C. Iyegbe, D. Blackwood, A. M. McIntosh, G. Kirov, D. Geschwind, R. M. Murray, M. Di Forti, E. Bramon, M. Gandal, C. M. Hultman, P. Sklar, INTERVAL Study, UK10K Consortium, A. Palotie, P. F. Sullivan, M.C. O’Donovan, M. J. Owen, J. C. Barrett, The contribution of rare variants to risk of schizophrenia in individuals with and without intellectual disability. Nat. Genet. 49, 1167–1173 (2017).

18. W. McLaren, L. Gil, S. E. Hunt, H. S. Riat, G. R. S. Ritchie, A. Thormann, P. Flicek, F. Cunningham, The Ensembl Variant Effect Predictor. Genome Biol. 17, 122 (2016).

19. T. Singh, T. Poterba, D. Curtis, H. Akil, M. Al Eissa, J. D. Barchas, N. Bass, T. B. Bigdeli, G. Breen, E. J. Bromet, P. F. Buckley, W. E. Bunney, J. Bybjerg-Grauholm, W. F. Byerley, S. B. Chapman, W. J. Chen, C. Churchhouse, N. Craddock, C. Curtis, C. M. Cusick, L. DeLisi, S. Dodge, M. A. Escamilla, S. Eskelinen, A. H. Fanous, S. V. Faraone, A. Fiorentino, L. Francioli, S. B. Gabriel, D. Gage, S. A. Gagliano Taliun, A. Ganna, G. Genovese, D. C. Glahn, J. Grove, M.-H. Hall, E. Hamalainen, H. O. Heyne, M. Holi, D. M. Hougaard, D. P. Howrigan, H. Huang, H.-G. Hwu, R. S. Kahn, H. M. Kang, K. Karczewski, G. Kirov, J. A. Knowles, F. S. Lee, D. S. Lehrer, F. Lescai, D. Malaspina, S. R. Marder, S. McCarroll, H. Medeiros, L. Milani, C. P. Morley, D. W. Morris, P. B. Mortensen, R. M. Myers, M. Nordentoft, N. L. O’Brien, A. M. Olivares, D. Ongur, W. H. Ouwehand, D. S. Palmer, T. Paunio, D. Quested, M. H. Rapaport, E. Rees, B. Rollins, F. Kyle Satterstrom, A. Schatzberg, E. Scolnick, L. Scott, S. I. Sharp, P. Sklar, J. W. Smoller, J. l. Sobell, M. Solomonson, C. R. Stevens, J. Suvisaari, G. Tiao, S. J. Watson, N. A. Watts, D. H. Blackwood, A. Borglum, B. M. Cohen, A. P. Corvin, T. Esko, N. B. Freimer, S. J. Glatt, C. M. Hultman, A. McQuillin, A. Palotie, C. N. Pato, M. T. Pato, A. E. Pulver, D. St. Clair, M. T. Tsuang, M. P. Vawter, J. T. Walters, T. Werge, R. A. Ophoff, P. F. Sullivan, M. J. Owen, M. Boehnke, M. O’Donovan, B. M. Neale, M. J. Daly, Exome sequencing identifies rare coding variants in 10 genes which confer substantial risk for schizophrenia. medRxiv, 2020.09.18.20192815 (2020).

20. F. Aguet, A. N. Barbeira, R. Bonazzola, A. Brown, S. E. Castel, B. Jo, S. Kasela, S. Kim-Hellmuth, Y. Liang, M. Oliva, P. E. Parsana, E. Flynn, L. Fresard, E. R. Gaamzon, A. R. Hamel, Y. He, F. Hormozdiari, P. Mohammadi, M. Muñoz-Aguirre, Y. Park, A. Saha, A. V. Segrć, B. J. Strober, X. Wen, V. Wucher, S. Das, D. Garrido-Martín, N. R. Gay, R. E. Handsaker, P. J. Hoffman, S. Kashin, A. Kwong, X. Li, D. MacArthur, J. M. Rouhana, M. Stephens, E. Todres, A. Viñuela, G. Wang, Y. Zou, C. D. Brown, N. Cox, E. Dermitzakis, B. E. Engelhardt, G. Getz, R. Guigo, S. B. Montgomery, B. E. Stranger, H. K. Im, A. Battle, K. G. Ardlie, T. Lappalainen, The GTEx Consortium, The GTEx Consortium atlas of genetic regulatory effects across human tissues,, doi:10.1101/787903.

21. H. K. Finucane, Y. A. Reshef, V. Anttila, K. Slowikowski, A. Gusev, A. Byrnes, S. Gazal, P.-R. Loh, C. Lareau, N. Shoresh, G. Genovese, A. Saunders, E. Macosko, S. Pollack, Brainstorm Consortium, J. R. B. Perry, J. D. Buenrostro, B. E. Bernstein, S. Raychaudhuri, S. A. McCarroll, B. M. Neale, A. L. Price, Heritability enrichment of specifically expressed genes identifies disease-relevant tissues and cell types. Nat. Genet. 50, 621–629 (2018).

22. D. P. Hibar, L. T. Westlye, T. G. M. van Erp, J. Rasmussen, C. D. Leonardo, J. Faskowitz, U. K. Haukvik, C. B. Hartberg, N. T. Doan, I. Agartz, A. M. Dale, O. Gruber, B. Krämer, S. Trost, B. Liberg, C. Abé, C. J. Ekman, M. Ingvar, M. Landén, S. C. Fears, N. B. Freimer, C. E. Bearden, Costa Rica/Colombia Consortium for Genetic Investigation of Bipolar Endophenotypes, E. Sprooten, D. C. Glahn, G. D. Pearlson, L. Emsell, J. Kenney, C. Scanlon, C. McDonald, D. M. Cannon, J. Almeida, A. Versace, X. Caseras, N. S. Lawrence, M. L. Phillips, D. Dima, G. Delvecchio, S. Frangou, T. D. Satterthwaite, D. Wolf, J. Houenou, C. Henry, U. F. Malt, E. Bøen, T. Elvsåshagen, A. H. Young, A. J. Lloyd, G. M. Goodwin, C. E. Mackay, C. Bourne, A. Bilderbeck, L. Abramovic, M. P. Boks, N. E. M. van Haren, R. A. Ophoff, R. S. Kahn, M. Bauer, A. Pfennig, M. Alda, T. Hajek, B. Mwangi, J. C. Soares, T. Nickson, R. Dimitrova, J. E. Sussmann, S. Hagenaars, H. C. Whalley, A. M. McIntosh, P. M. Thompson, O. A. Andreassen, Subcortical volumetric abnormalities in bipolar disorder. Mol. Psychiatry. 21, 1710–1716 (2016).

23. A. Ganna, G. Genovese, D. P. Howrigan, A. Byrnes, M. Kurki, S. M. Zekavat, C. W. Whelan, M. Kals, M. G. Nivard, A. Bloemendal, J. M. Bloom, J. I. Goldstein, T. Poterba, C. Seed, R. E. Handsaker, P. Natarajan, R. Mägi, D. Gage, E. B. Robinson, A. Metspalu, V. Salomaa, J. Suvisaari, S. M. Purcell, P. Sklar, S. Kathiresan, M. J. Daly, S. A. McCarroll, P. F. Sullivan, A. Palotie, T. Esko, C. Hultman, B. M. Neale, Ultra-rare disruptive and damaging mutations influence educational attainment in the general population. Nat. Neurosci. 19, 1563–1565 (2016).

24. J. Cotney, R. A. Muhle, S. J. Sanders, L. Liu, A. J. Willsey, W. Niu, W. Liu, L. Klei, J. Lei, J. Yin, S. K. Reilly, A. T. Tebbenkamp, C. Bichsel, M. Pletikos, N. Sestan, K. Roeder, M. W. State, B. Devlin, J. P. Noonan, The autism-associated chromatin modifier CHD8 regulates other autism risk genes during human neurodevelopment. Nat. Commun. 6, 6404 (2015).

25. S. M. Weyn-Vanhentenryck, A. Mele, Q. Yan, S. Sun, N. Farny, Z. Zhang, C. Xue, M. Herre, P. A. Silver, M. Q. Zhang, A. R. Krainer, R. B. Darnell, C. Zhang, HITS-CLIP and integrative modeling define the Rbfox splicing-regulatory network linked to brain development and autism. Cell Rep. 6, 1139–1152 (2014).

26. S. J. Sanders, X. He, A. J. Willsey, A. G. Ercan-Sencicek, K. E. Samocha, A. E. Cicek, M. T. Murtha, V. H. Bal, S. L. Bishop, S. Dong, A. P. Goldberg, C. Jinlu, J. F. Keaney 3rd, L. Klei, J. D. Mandell, D. Moreno-De-Luca, C. S. Poultney, E. B. Robinson, L. Smith, T. Solli-Nowlan, M. Y. Su, N. A. Teran, M. F. Walker, D. M. Werling, A. L. Beaudet, R. M. Cantor, E. Fombonne, D. H. Geschwind, D. E. Grice, C. Lord, J. K. Lowe, S. M. Mane, D. M. Martin, E. M. Morrow, M. E. Talkowski, J. S. Sutcliffe, C. A. Walsh, T. W. Yu, Autism Sequencing Consortium, D. H. Ledbetter, C. L. Martin, E. H. Cook, J. D. Buxbaum, M. J. Daly, B. Devlin, K. Roeder, M. W. State, Insights into Autism Spectrum Disorder Genomic Architecture and Biology from 71 Risk Loci. Neuron. 87, 1215–1233 (2015).

27. Bipolar Disorder and Schizophrenia Working Group of the Psychiatric Genomics Consortium. Electronic address, Bipolar Disorder and Schizophrenia Working Group of the Psychiatric Genomics Consortium, Genomic Dissection of Bipolar Disorder and Schizophrenia, Including 28 Subphenotypes. Cell. 173, 1705–1715.e16 (2018).

28. K. J. Karczewski, L. C. Francioli, G. Tiao, B. B. Cummings, J. Alföldi, Q. Wang, R. L. Collins, K. M. Laricchia, A. Ganna, D. P. Birnbaum, L. D. Gauthier, H. Brand, M. Solomonson, N. A. Watts, D. Rhodes, M. Singer-Berk, E. M. England, E. G. Seaby, J. A. Kosmicki, R. K. Walters, K. Tashman, Y. Farjoun, E. Banks, T. Poterba, A. Wang, C. Seed, N. Whiffin, J. X. Chong, K. E. Samocha, E. Pierce-Hoffman, Z. Zappala, A. H. O’Donnell-Luria, E. V. Minikel, B. Weisburd, M. Lek, J. S. Ware, C. Vittal, I. M. Armean, L. Bergelson, K. Cibulskis, K. M. Connolly, M. Covarrubias, S. Donnelly, S. Ferriera, S. Gabriel, J. Gentry, N. Gupta, T. Jeandet, D. Kaplan, C. Llanwarne, R. Munshi, S. Novod, N. Petrillo, D. Roazen, V. Ruano-Rubio, A. Saltzman, M. Schleicher, J. Soto, K. Tibbetts, C. Tolonen, G. Wade, M. E. Talkowski, Genome Aggregation Database Consortium, B. M. Neale, M. J. Daly, D. G. MacArthur, The mutational constraint spectrum quantified from variation in 141,456 humans. Nature. 581, 434–443 (2020).

29. D. P. Howrigan, S. A. Rose, K. E. Samocha, M. Fromer, F. Cerrato, W. J. Chen, C. Churchhouse, K. Chambert, S. D. Chandler, M. J. Daly, A. Dumont, G. Genovese, H.-G. Hwu, N. Laird, J. A. Kosmicki, J. L. Moran, C. Roe, T. Singh, S.-H. Wang, S. V. Faraone, S. J. Glatt, S. A. McCarroll, M. Tsuang, B. M. Neale, Exome sequencing in schizophrenia-affected parent-offspring trios reveals risk conferred by protein-coding de novo mutations. Nat. Neurosci. 23, 185–193 (2020).

30. L. Freland, J.-M. Beaulieu, Inhibition of GSK3 by lithium, from single molecules to signaling networks. Front. Mol. Neurosci. 5, 14 (2012).

31. B. K. Kishore, C. M. Ecelbarger, Lithium: a versatile tool for understanding renal physiology. Am. J. Physiol. Renal Physiol. 304, F1139–49 (2013).

32. R. S. Jope, Lithium and GSK-3: one inhibitor, two inhibitory actions, multiple outcomes. Trends Pharmacol. Sci. 24, 441–443 (2003).

33. H. V. Firth, S. M. Richards, A. P. Bevan, S. Clayton, M. Corpas, D. Rajan, S. Van Vooren, Y. Moreau, R. M. Pettett, N. P. Carter, DECIPHER: Database of Chromosomal Imbalance and Phenotype in Humans Using Ensembl Resources. Am. J. Hum. Genet. 84, 524–533 (2009).

34. F. K. Satterstrom, J. A. Kosmicki, J. Wang, M. S. Breen, S. De Rubeis, J.-Y. An, M. Peng, R. Collins, J. Grove, L. Klei, C. Stevens, J. Reichert, M. S. Mulhern, M. Artomov, S. Gerges, B. Sheppard, X. Xu, A. Bhaduri, U. Norman, H. Brand, G. Schwartz, R. Nguyen, E. E. Guerrero, C. Dias, Autism Sequencing Consortium, iPSYCH-Broad Consortium, C. Betancur, E. H. Cook, L. Gallagher, M. Gill, J. S. Sutcliffe, A. Thurm, M. E. Zwick, A. D. Børglum, M. W. State, A. E. Cicek, M. E. Talkowski, D. J. Cutler, B. Devlin, S. J. Sanders, K. Roeder, M. J. Daly, J. D. Buxbaum, Large-Scale Exome Sequencing Study Implicates Both Developmental and Functional Changes in the Neurobiology of Autism. Cell. 180, 568–584.e23 (2020).

35. Epi25 Collaborative. Electronic address, Epi25 Collaborative, Ultra-Rare Genetic Variation in the Epilepsies: A Whole-Exome Sequencing Study of 17,606 Individuals. Am. J. Hum. Genet. 105, 267–282 (2019).

36. C. Tanji, H. Yamamoto, N. Yorioka, N. Kohno, K. Kikuchi, A. Kikuchi, A-kinase anchoring protein AKAP220 binds to glycogen synthase kinase-3beta (GSK-3beta) and mediates protein kinase A-dependent inhibition of GSK-3beta. J. Biol. Chem. 277, 36955–36961 (2002).

37. E. Beurel, S. F. Grieco, R. S. Jope, Glycogen synthase kinase-3 (GSK3): regulation, actions, and diseases. Pharmacol. Ther. 148, 114–131 (2015).

