## Supplementary materials for "Exome sequencing in bipolar disorder reveals shared risk gene *AKAP11* with schizophrenia"

### Exome sequencing in bipolar disorder reveal shared risk gene *AKAP11* with schizophrenia: Supplementary Materials

1. Analytic and Translational Genetics Unit, Department of Medicine, Massachusetts General Hospital, Boston, Massachusetts, USA
2. Stanley Center for Psychiatric Research, Broad Institute of MIT and Harvard, Cambridge, Massachusetts, USA
3. Department of Clinical Sciences, Psychiatry, Umea University, Umea, Sweden
4. Division of Psychiatry, University College London, London, UK
5. University of Edinburgh, Edinburgh, UK
6. Department of Psychiatry, UMC Utrecht, Utrecht, The Netherlands
7. Biogen, Cambridge, Massachusetts, USA
8. Program in Medical and Population Genetics, Broad Institute of Harvard and MIT, Cambridge, Massachusetts, USA
9. Trinity College Dublin, Dublin, Ireland
10. National Centre for Mental Health, Division of Psychiatry and Clinical Neurosciences, Cardiff University, Cardiff, UK
11. UCL Genetics Institute, University College London, London, UK
12. Centre for Psychiatry, Queen Mary University of London, London, UK
13. Division of Psychological Medicine and Clinical Neurosciences, Cardiff University, Cardiff, UK
14. Sheppard Pratt, 6501 North Charles Street, Baltimore, Maryland, USA
15. Department of Psychiatry and Behavioral Sciences, Johns Hopkins School of Medicine, Baltimore, Maryland, USA
16. Weill Institute for Neurosciences, University of California, San Francisco, California, USA
17. Department of Psychological Medicine, University of Worcester, Worcester, UK
18. Department of Psychiatry and Neurochemistry, Institute of Neuroscience and Physiology, Sahlgrenska Academy, University of Gothenburg, Gothenburg, Sweden
19. Department of Pharmacology, Institute of Neuroscience and Physiology, Sahlgrenska Academy, University of Gothenburg, Gothenburg, Sweden
20. Division of Psychiatry, Icahn School of Medicine at Mount Sinai, New York, New York, USA
21. Department of Medical Epidemiology and Biostatistics, Karolinska Institutet, Stockholm, Sweden
22. Division of Genomics & Bioinformatics, Washington University School of Medicine, St. Louis, Michigan, USA
23. National University of Ireland, Galway, Ireland
24. MRC Centre for Neuropsychiatric Genetics and Genomics, Division of Psychological Medicine and Clinical Neurosciences, Cardiff University, Cardiff, UK
25. Center for Neurobehavioral Genetics, University of California, Los Angeles, California, USA
26. Department of Psychiatry, Erasmus Medical Center, Erasmus University, Rotterdam, The Netherlands
27. Vrije Universiteit, Amsterdam, The Netherlands
28. Department of Psychiatry, Psychosomatic Medicine and Psychiatry, University Hospital Frankfurt - Goethe University, Frankfurt, Germany
29. Institute for Human Genetics, University of California, San Francisco, San Francisco, California, USA
30. Division of Research, Kaiser Permanente Northern California, Oakland, California, USA
31. Biostatistics and Center for Statistical Genetics, University of Michigan, Ann Arbor, Michigan, USA
32. Psychiatric and Neurodevelopmental Genetics Unit, Massachusetts General Hospital, Boston, Massachusetts, USA
33. Department of Psychiatry, Harvard Medical School, Boston, Massachusetts, USA
34. University of Aberdeen, Aberdeen, UK
35. Division of Psychiatric Genomics, Icahn School of Medicine at Mount Sinai, New York, New York, USA
36. Stanley Division of Developmental Neurovirology, Johns Hopkins University, Baltimore, Maryland, USA

### Sample collections

The BipEx cohort aggregates 13 separate sample collections, involving 21 primary investigators across 6 separate countries. We performed careful quality control steps to variants and samples, detailed in the Exome QC section. The aggregated dataset consists of 39,617 individuals, 16,486 of which have been diagnosed with bipolar disorder, and 17,213 with no known psychiatric diagnosis. Of the remaining individuals, 5,483 have a schizophrenia diagnosis (which we use as positive controls in the rare variant burden analyses), 87 have a separate psychiatric diagnosis, and 32 lack phenotypic information. Full details of PI sample contributions prior to curation of sequence data are provided in Table S1. Following curation, the case and control count is displayed in Table S4. Breakdown of bipolar cases with age of onset information for age of first impairment is provided in Table S6, and a breakdown of sample sizes with psychosis information is displayed in Table S7. Sample collection and phenotype tables are also available at https://astheeggeggs.github.io/BipEx/qc.html.

| PI | Location | BD | BD1 | BD2 | BDNOS | SAD | SCZ | Other | Unknown | Controls | Total |
|---|---|---|---|---|---|---|---|---|---|---|---|
| Andreas Reif | Wurzburg, GER | 7 | 216 | 159 | 15 | 0 | 0 | 0 | 14 | 414 | 825 |
| Andrew McQuillin<br>Hugh Gurling | London, UK | 228 | 1,309 | 372 | 0 | 157 | 1,595 | 0 | 0 | 1,203 | 4,864 |
| Robert Yolken<br>Faith Dickerson | Baltimore, USA | 8 | 117 | 9 | 5 | 0 | 0 | 8 | 0 | 126 | 273 |
| Danielle Posthuma | Amsterdam, NED | 0 | 0 | 0 | 0 | 0 | 0 | 0 | 1 | 948 | 949 |
| David St Clair | Aberdeen, UK | 0 | 0 | 0 | 0 | 0 | 564 | 0 | 1 | 331 | 896 |
| Derek Morris<br>Aiden Corvin | Dublin, IRE | 0 | 180 | 0 | 0 | 11 | 29 | 3 | 0 | 9 | 232 |
| Douglas Blackwood | Edinburgh, UK | 401 | 368 | 114 | 2 | 6 | 304 | 0 | 0 | 64 | 1,259 |
| Fernando Goes | Baltimore, USA | 0 | 241 | 0 | 0 | 0 | 0 | 0 | 0 | 0 | 241 |
| Jordan Smoller | Boston, USA | 361 | 2,122 | 390 | 576 | 52 | 0 | 0 | 0 | 3,498 | 6,999 |
| Michael O Donovan | Cardiff, UK | 0 | 0 | 0 | 0 | 11 | 2,986 | 1 | 0 | 0 | 2,998 |
| Michael Owen | Cardiff, UK | 0 | 0 | 0 | 0 | 0 | 0 | 0 | 0 | 1,106 | 1,106 |
| Mikael Landén | Stockholm, SWE | 138 | 2,364 | 1,753 | 905 | 1 | 0 | 0 | 0 | 761 | 5,922 |
| Nancy Pedersen | Stockholm, SWE | 0 | 0 | 0 | 0 | 0 | 0 | 0 | 0 | 4,780 | 4,780 |
| Nick Craddock<br>Arianna Di Florio<br>Ian Jones<br>Lisa Jones<br>James Walters | Cardiff, UK | 85 | 1,518 | 772 | 67 | 57 | 4 | 17 | 0 | 0 | 2,520 |
| Roel Ophoff | Utrecht, NED | 1 | 1,032 | 169 | 10 | 21 | 1 | 58 | 16 | 663 | 1,971 |
| Rolf Adolfsson | Umea, SWE | 0 | 320 | 149 | 3 | 0 | 0 | 0 | 0 | 459 | 931 |
| Willem Ouwehand | Cambridge, UK | 0 | 0 | 0 | 0 | 0 | 0 | 0 | 0 | 2,851 | 2,851 |
| **Total** | | **1,229** | **9,787** | **3,887** | **1,583** | **316** | **5,483** | **87** | **32** | **17,213** | **39,617** |

**Table S1**: Detailed summary of subtype sample contributions across PIs and geographies. BD=BD without a fine subclassification, BD1=bipolar I disorder, BD2=bipolar II disorder, BDNOS=bipolar disorder not otherwise specified, SAD=schizoaffective disorder, BD+SAD=bipolar disorder and schizoaffective disorder combined, SCZ=schizophrenia, other=other unspecified case, unknown=unknown case status.

### Cohort descriptions and bipolar subtype definitions

**Aberdeen, UK**

***PI: David St Clair***

All participants self-identified as born in the British Isles (95% in Scotland). All schizophrenia cases met the DSM-IV (*1*) and International Classification of Diseases 10th edition (ICD-10) (*2*) criteria for schizophrenia. Diagnosis was made by Operational Criteria Checklist (OPCRIT) (*3*, *4*). All case participants were outpatients or stable in-patients. Detailed medical and psychiatric histories were collected. Controls were volunteers recruited through general practices in Scotland. Practice lists were screened for potentially suitable volunteers by age and sex and by exclusion of subjects with major mental illness or use of neuroleptic medication. Volunteers who replied to a written invitation were interviewed using a short questionnaire to exclude major mental illness in individuals themselves and first degree relatives. All cases and controls gave informed consent. The study was approved by both local and multiregional academic ethical committees.

**Amsterdam, NED**

***PI: Danielle Posthuma***

Controls taken from the NESCOG study, described previously (*5*). NESCOG contains both a general population and family-based sample of which closely related individuals were excluded. Data were collected on cognitive tasks, behavioral conditions, life events, personality and environmental factors. To correct for undiagnosed attention deficit hyperactivity disorder (ADHD) status, participants scoring over three standard deviations above the mean on the Conners' Adult ADHD Rating Scale (CAARS) (*6*), or the Attention Problems scale of the Young Adult Self Report (YASR) (*7*) were excluded. To correct for autism spectrum disorder (ASD) status, participants scoring over three standard deviations above the mean on the Autism Quotient (AQ) (*8*) were removed.

**Baltimore, USA**

***PIs: Faith Dickerson, Robert Yolken***

Samples were collected as part of a larger study about infectious agents and immune factors in serious mental illness. Psychiatric participants were recruited at a large psychiatric health system and non-psychiatric controls from the same geographic region. The diagnosis of psychiatric and non-psychiatric participants was confirmed with a structured clinical interview (*9*, *10*). All participants provided written informed consent. The study was approved by the IRB of the institution where the study was performed and included a data sharing agreement.

***PI: Fernando Goes***

Cases represented independent probands from a European-American family sample that was collected at Johns Hopkins University from 1988-2010. Families had at least 2 additional relatives with a major mood disorder (defined as bipolar disorder type 1, bipolar type 2 or recurrent major depressive disorder). Diagnostic interviews were performed using the Schedule for Affective Disorders and Schizophrenia-Lifetime Version and the Diagnostic Instrument for Genetics Studies. All cases underwent best-estimate diagnostic procedures. Diagnoses were based on DSM-III and DSM-IV (*1*) criteria. Probands from this sample have been previously studied in family based linkage and exome studies (*11*, *12*).

**Boston, USA**

***PI: Jordan Smoller***

Cases and controls were collected as part of the International Cohort Collection for Bipolar Disorder (ICCBD), (*13*, *14*). The Massachusetts General Hospital site of the ICCBD collected DNA from cases and controls by linking discarded blood samples to de-identified electronic health record (EHR) data. Cases and controls were identified by deriving EHR-based phenotyping algorithms applied to the Partners Healthcare Research Patient Data Registry (RPDR), described in detail previously (*15*). Bipolar subtypes were defined by Diagnostic and Statistical Manual of Mental Disorders-IV (DSM-IV) (*1*). Regular expression rules were used to extract mention by clinician in an inpatient or outpatient note or ICD-9/DSM-IV (*1*, *16*, *17*) code indicating Bipolar type. Full details of the algorithm are provided in (*14*).

**Cambridge, UK**

***PI: Willem Ouwehand***

Controls taken from the Wellcome Trust case control consortium (WTCCC) described elsewhere (*18*).

**Cardiff, UK**

***PIs: Nick Craddock, Arianna Di Florio, Ian Jones , Lisa Jones, James Walters***

Cases were all over the age of 17 years, living in the UK and of European descent. Cases were recruited via systematic and not systematic methods as part of the Bipolar Disorder Research Network project (www.bdrn.org), provided written informed consent and were interviewed using a semi-structured diagnostic interview, the Schedules for Clinical Assessment in Neuropsychiatry. Based on the information gathered from the interview and case notes review, best-estimate lifetime diagnosis was made according to DSM-IV. Inter-rater reliability was formally assessed using 20 randomly selected cases (mean κ Statistic = 0.85). In the current study we included cases with a lifetime diagnosis of DSM-IV bipolar disorder or schizo-affective disorder, bipolar type. The BDRN study has UK National Health Service (NHS) Research Ethics Committee approval and local Research and Development approval in all participating NHS Trusts/Health Boards. All subjects gave written informed consent.

***PIs: Michael O Donovan, Michael Owen***

The schizophrenia case sample included European ancestry schizophrenia cases recruited in the British Isles and has been described previously (*19*). All cases gave written informed consent to. The study was approved by the Multicentre Research Ethics Committee in Wales and Local Research Ethics Committees from all participating sites. The control sample used the WTCCC sample described elsewhere, (*18*) but included similar numbers of individuals from the 1958 British Birth Cohort and a panel of consenting blood donors (UK Blood Service). Additional controls, held by Cardiff University, were recruited from the UK National Blood Transfusion Service. They were not specifically screened for psychiatric illness. All control samples were from participants who provided informed consent (*20*).

**Dublin, IRE**

***PIs: Derek Morris, Aiden Corvin***

Samples were collected as part of a larger study of the genetics of psychotic disorders in the Republic of Ireland, under protocols approved by the relevant IRBs and with written informed consent that permitted repository use. Cases were recruited from Hospitals and Community psychiatric facilities in Ireland by a psychiatrist or psychiatric nurse trained to use the SCID (*21*, *22*). Diagnosis was based on the structured interview supplemented by case note review and collateral history where available. All diagnoses were reviewed by an independent reviewer. Controls were ascertained with informed consent from the Irish GeneBank and represented blood donors who met the same ethnicity criteria as cases. Controls were not specifically screened for psychiatric illness.

**Edinburgh, UK**

***PI: Douglas Blackwood***

This sample comprised Caucasian individuals contacted through the inpatient and outpatient services of hospitals in South East Scotland. A BD1 diagnosis was based on an interview with the patient using the SADS-L supplemented by case note review and frequently by information from medical staff, relatives and caregivers. Final diagnoses, based on DSM-IV criteria were reached by consensus between two trained psychiatrists. Ethnically-matched controls from the same region were recruited through the South of Scotland Blood Transfusion Service. Controls were not directly screened to exclude those with a personal or family history of psychiatric illness. The study was approved by the Multi-Centre Research Ethics Committee for Scotland and patients gave written informed consent for the collection of DNA samples for use in genetic studies.

**London, UK**

***PIs: Andrew McQuillin, Hugh Gurling***

The UCL sample comprised Caucasian individuals who were ascertained and received clinical diagnoses of bipolar 1 disorder according to UK National Health Service (NHS) psychiatrists at interview using the categories of ICD10. In addition bipolar subjects were included only if both parents were of English, Irish, Welsh or Scottish descent and if three out of four grandparents were of the same descent. All volunteers read an information sheet approved by the Metropolitan Medical Research Ethics Committee who also approved the project for all NHS hospitals. Written informed consent was obtained from each volunteer. The UCL control subjects were recruited from London branches of the National Blood Service, from local NHS family doctor clinics and from university student volunteers. All control subjects were interviewed with the SADS-L to exclude all psychiatric disorders.

**Stockholm, SWE**

***PI: Mikael Landén***

**SWEBIC (Swedish Bipolar Cohort Collection), SWE**

Data in SWEBIC combines phenotypic data from three routes of collection:

1. The St. Göran Bipolar Project cohort (**SBP**).
2. SWEBIC samples recruited from the Swedish National quality assurance register for bipolar disorders (**BipoläR**).
3. SWEBIC samples recruited from the Swedish Hospital Discharge Register (**HDR**).

**SBP:** DSM-IV-criteria was evaluated by psychiatrists or residents in psychiatry using a Swedish version of the Affective Disorder Evaluation (ADE) employed in the Systematic Treatment Enhancement Program for Bipolar Disorder (STEP-BD) study (*21*) which includes the Structured Clinical Interview for DSM Disorders (SCID) (*21*, *22*) module for affective disorders.
**BipoläR:** Diagnostic phenotyping was made by registering physicians in the QA-register according to DSM-IV criteria.
**HDR**: Bipolar disorder cases were selected based on a validated algorithm using ICD-codes with a positive predictive value of 0.92 (*23*) Bipolar subdiagnoses (BD1, BD2, BDNOS) were made by trained research nurses using a structured telephone interview.

***PI: Nancy Pederson***

Controls were sourced from the LifeGene Biorepository at the Karolinska Institute, described in detail previously (*24*).

**Umea, SWE**

***PI: Rolf Adolfsson***

Bipolar disorder outpatients at the Affective Unit at the Psychiatric Clinic at the University Hospital (Umeå, Sweden) were enrolled in this study between 1998 and 2007 (*25*). Patients were characterised clinically in a number of ways, including the MINI (*26*), the Family Interview for Genetic Studies (FIGS), the Diagnostic Interview for Genetic Studies (DIGS) (*27*), and the Schedules for Clinical Assessment in Neuropsychiatry (SCAN) (*28*). Final subtype diagnoses were evaluated in line with the DSM-IV-TR (*29*) and determined through consensus of two research psychiatrists. Controls in the data set were a randomly sampled subset of the ‘Betula study’ chosen to be representative of the population of the region.

**Utrecht, NED**

***PI: Roel Ophoff***

The case sample consisted of inpatients and outpatients recruited through psychiatric hospitals and institutions throughout the Netherlands. Cases with DSM-IV bipolar disorder, determined after interview with the SCID (*21*, *22*) were included in the analysis. Controls were collected in parallel at different sites in the Netherlands and were volunteers with no psychiatric history after screening with the Mini-International Neuropsychiatric Interview (MINI) (*26*). Ethical approval was provided by UCLA, the University Medical Center Utrecht, and local ethics committees and all participants gave written informed consent.

**Wurzburg, GER**

***PI: Andreas Reif***

Cases were recruited from consecutive admissions to psychiatric in-patient units at the University Hospital Würzburg. All cases received a lifetime diagnosis of BD according to the DSM-IV criteria using a consensus best-estimate procedure based on all available information, including semi-structured diagnostic interviews using the Association for Methodology and Documentation in Psychiatry (*29*, *30*), medical records and the family history. In addition, the OPCRIT (*3*, *4*) system was used for the detailed polydiagnostic documentation of symptoms. Control subjects were healthy participants who were recruited from the community of the same region as cases (*31*). They were of Caucasian descent and fluent in German. Exclusion criteria were manifest or lifetime DSM-IV axis I disorder, severe medical conditions, intake of psychoactive medication as well as alcohol abuse or abuse of illicit drugs. Absence of DSM-IV axis I disorder was ascertained using the German versions of the MINI (*26*). IQ was above 85 as ascertained by the German version of the Culture Fair Intelligence Test 2. Study protocols were reviewed and approved by the ethical committee of the Medical Faculty of the University of Würzburg. All subjects provided written informed consent.

### Sequence data production

#### Exome Sequencing and Alignment

Exome sequencing was performed at the Broad Institute of Harvard and MIT from July 2017 to September 2018. Processing included sample QC using the picogreen assay to measure for sample volume, concentration and DNA yield. Sample library preparation was carried out using Illumina Nextera, followed by hybrid capture using Illumina rapid capture enrichment of 37Mb target. Sequencing was performed on HiSeqX instruments to 150bp paired reads. Sample identification checking was carried out to confirm all samples. Sequencing was run until hybrid selection libraries met or exceeded 85% of targets at 20x, comparable to ~55x mean coverage. Data delivery per sample includes a demultiplexed, aggregated into a BAM file and processed through a pipeline based on the Picard suite of software tools. The BWA aligner mapped reads onto the human genome build 38 (GRCh38). Single nucleotide polymorphism and insertions/deletions were joint called across all samples using Genome Analysis Toolkit (GATK) (*32*) HaplotypeCaller package version 4.0.10 to produce a version 4.2 variant callset file (VCF). Variant call accuracy was estimated using the GATK Variant Quality Score Recalibration (VQSR) approach (*33*).

### Exome Quality Control

Throughout, to perform quality control and a subset of the downstream analyses, we made use of Hail, an open-source, general-purpose, Python-based data analysis library with a particular focus on the analysis of large-scale genetic data (website: https://www.hail.is, GitHub: https://github.com/hail-is/hail). We made use of the general purpose hail framework to create our own methods, as well as take advantage of the functionality that has been rewritten to enable fast and scalable analysis of large exome and genome sequencing projects. Unless otherwise stated, all of the data curation and quality control steps were performed in hail 0.2.
Briefly, we perform a series of hard-filters on genotype and variant metrics (Table S3), followed by a collection of hard-filters on sample metrics (Table S4). We confirm genotype sex with reported sex, remove related individuals, and restrict analysis to samples of continental European ancestry where we have suffucient sampl size and balanced case-control counts (Table S4). Finally we filter based on a second collection of sample and variant hard-filters (Tables S3-4).

#### Initial Hard Filters

A series of quality control (QC) steps were run to clean and curate the sequence data. We first apply a collection of genotype filters, removing genotypes according to the following criteria: If homozygous reference, remove if at least one of the following is true: phred-scaled genotype quality (GQ) < 20, depth (DP) < 10. If heterozygous, remove if at least one of the following is true: (reference allele depth + alternative allele depth) divided by total depth < 0.8, alternative allele depth divided by total depth < 0.2, reference genotype quality < 20, depth < 10. If homozygous alternate, remove if at least one of the following is true: alternative allele depth divided by total depth < 0.8, reference genotype quality < 20, depth < 10. We then apply a series of initial variant filtering steps: removing sites with more than 6 alleles, that fail variant quality score recalibration (VQSR) (*33*)(*32*); (*33*), lie within a low complexity region (LCR) of the genome (*34*), or fall over 50 base pairs outside the ICE exome sequencing target intervals. In addition, we perform a series of empirically derived genotype call rate filters (set at 0.97) and remove sites that become invariant after applying this filter (Table S3). As an initial pass to remove low quality and contaminated samples, we filter out samples with call rate < 0.93, free-mix contamination > 0.02 (*35*), chimeric read percentage > 0.015, mean read depth < 30x or mean genotype quality < 55 (Table S4).

Empirically derived hard filters to filter on these metrics are detailed in Table S3. In each plot, jittered scatters display the distribution for each sequencing batch, coloured according to sample collection. Boxplots behind the scatter display the median and interquartile range for each sequencing batch.

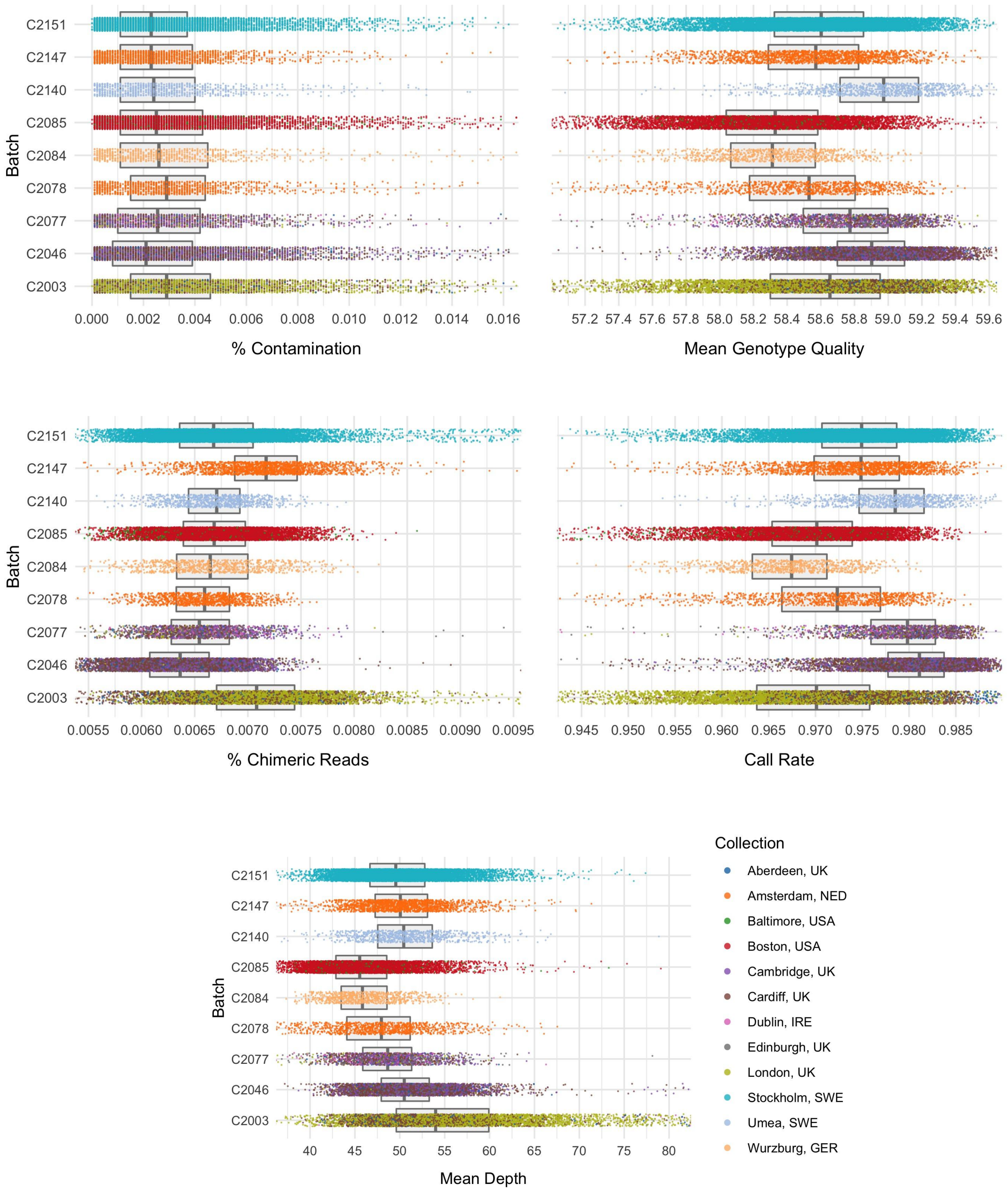


**Figure S1:** Distributions of variant metrics following restriction to variants passing VQSR, lying outside low-complexity regions and inside the padded (50bp) ICE target intervals, and prior to the initial hard sample filters (call rate > 0.93, FREEMIX contamination (%) < 0.02, percentage chimeras < 0.015, mean depth > 30, mean genotype quality > 55).

#### Sex Imputation and Relatedness

To confirm participant sex, filter out related samples, and calculate principal components, we extracted high quality common variants (allele frequency between 0.01 to 0.99 with high call rate (> 0.98)) and LD-prune to pseudo-independent SNPs using `--indep 50 5 2` in PLINK (*36*, *37*). Using autosomal markers (49,366 SNPs), we determine relatedness within the sample, and iteratively prune out samples until no pair exhibited $\hat{\pi} > 0.2$ to ensure that first and second degree relatives are filtered out. When reported sex does not match genotyped sex, it may signal potential sample swaps in the data. Using the *F*-statistic for each sample using the subset of the non-pseudo autosomal region on chromosome X (1275 SNPs), we identify and remove samples where reported sex information is not confirmed in the sequence data (Figure S2).

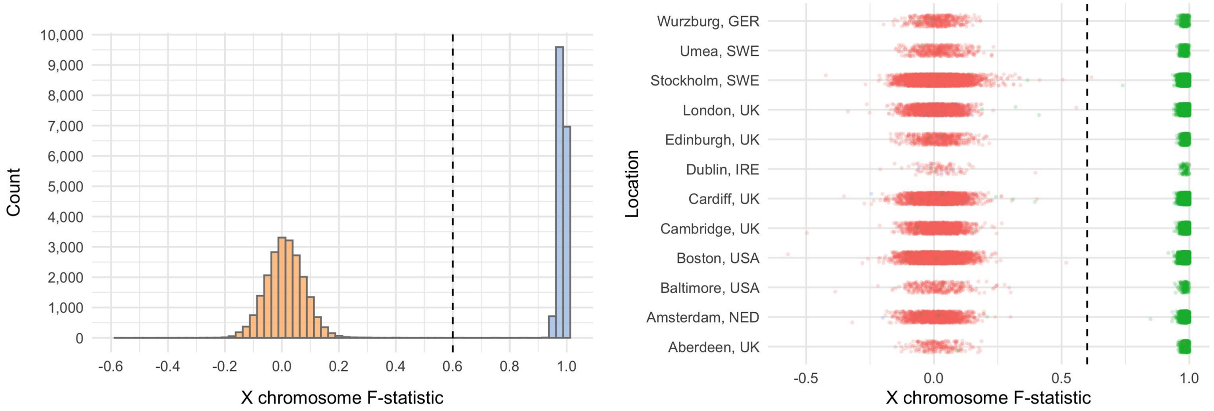


**Figure S2:** Histogram and scatterplots of X chromosome *F*-statistic by collection. Samples lying to the left and right of the dashed line were called as female and male respectively.

#### Principal component based population inference

We then merged the remaining samples with the 1000 Genomes phase 3 dataset (*38*), and computed principal components (PCs) using the LD pruned autosomal variants (49,366 SNPs). To ensure adequate case-control matching, we removed samples outside of the continental European population (EUR) using a random forest classifier trained on the EUR subset of 1000 Genomes (Figure S3), retaining samples with probability > 0.95 of being European according to the classifier. Additionally, we removed Ashkenazi Jewish samples by running principal components analysis (PCA) on samples recruited in the United States and identifying a distinct Ashkenazi Jewish cluster. Using this labelling, we trained another random forest classifier and removed additional Ashkenazi Jewish samples from downstream analysis, again using a hard cutoff of probability > 0.95 of belonging to the main European cluster (Table S3).

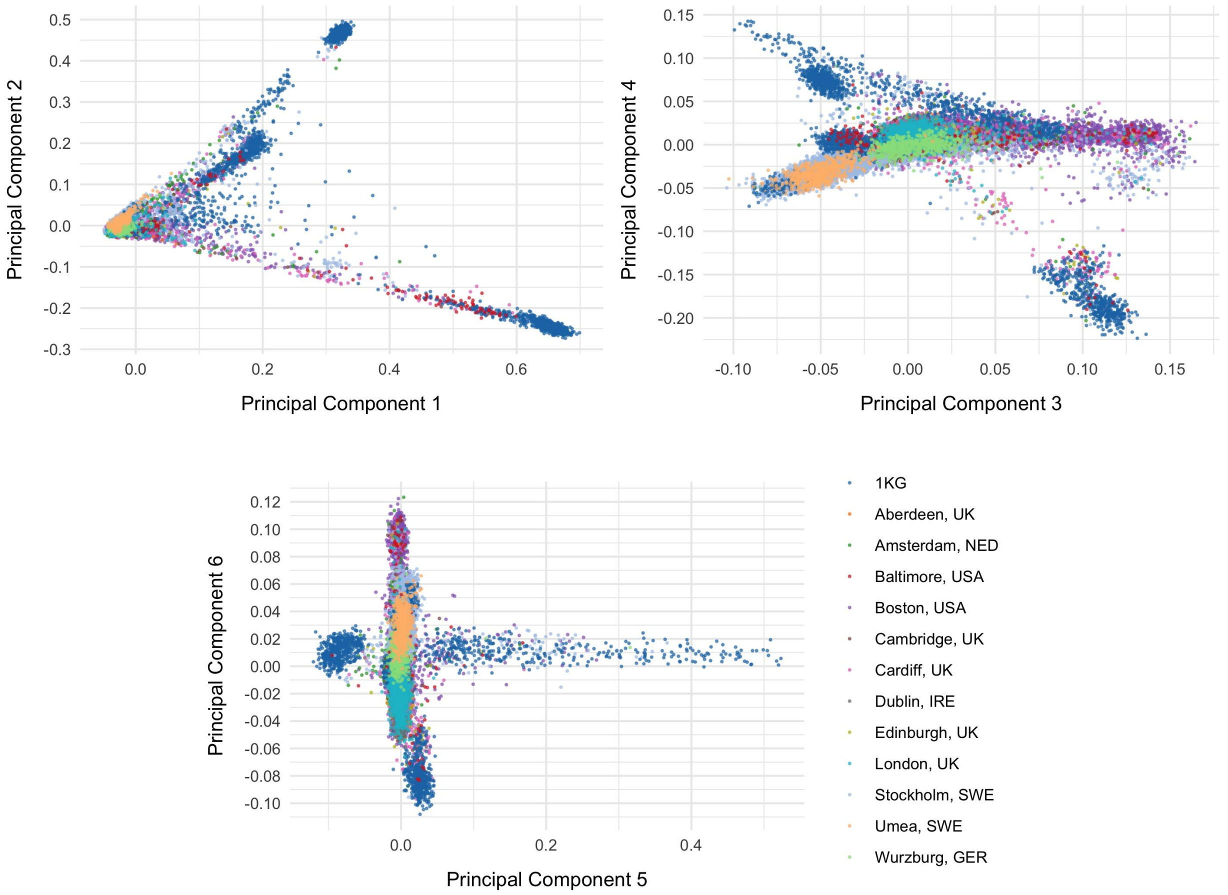


**Figure S3:** Scatterplots of principal components of BipEx samples together with 1000 Genomes samples. Points are coloured according to sample collection, with 1000 Genomes samples coloured in blue. 1000 Genomes super-populations labels were used to train a random forest classifier.

##### Final Hard Filters

For our second round of variant and sample filtering, we filter out variants based on call rate (BD call rate < 0.97, control call rate < 0.97, overall call rate < 0.97), difference in call rate between bipolar cases and controls (> 0.02), and remove variants not in Hardy-Weinberg equilibrium ($p < 10^{-6}$); Table S3. After restricting to these high quality variants (Figure S4), we perform a final set of sample filters to finalise the QCed data. We evaluate a collection of sample metrics and remove samples falling outside three standard deviations of the sequencing batch mean (Ti/Tv, Het/HomVar, Insertion/Deletion ratios) or cohort location (as defined by recruitment centre; *n* singletons) mean (Table S4, Figure S4). The resultant dataset consisting of 33,527 samples across 12 locations in Europe and the United States is summarised in Table S1. Following our QC pipeline, average heterozygote allele balance was 0.484, with 1.52% of samples lying below 0.3, and Ti/Tv became comparable between sequencing batches and sample collection. Further, average sample Ti/Tv within the targeted exome region was ~3.1 (rather than the 50bp padded), in line with expectation for populations of European ancestry (Figure S5).

Full details, code and all files required to run our pipeline are available at github.com/astheeggeggs/BipEx.

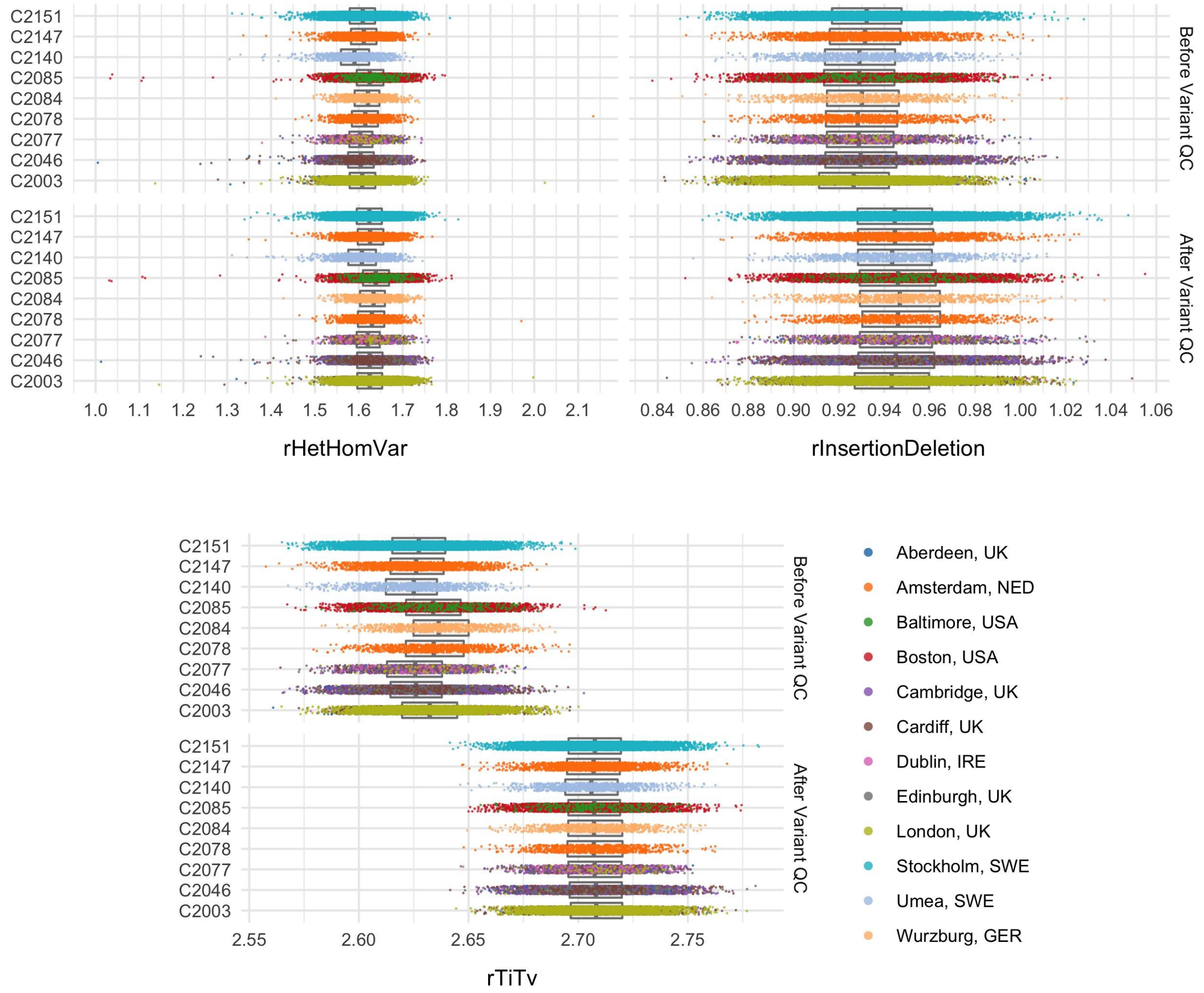


**Figure S4:** Distributions of variant metrics before and after the second set of empirically derived hard variant filters (BD call rate > 0.97, control call rate > 0.97, overall call rate > 0.97), difference in call rate between bipolar cases and controls (< 0.02), and remove variants not in Hardy-Weinberg equilibrium ($p > 10^{-6}$). In each plot, jittered scatters display the distribution for each sequencing batch, coloured according to sample collection. Boxplots behind the scatter display the median and interquartile range for each sequencing batch. Points shown are following variants hard-filters and prior to removal of variants with metrics outside 3 sds of the sequencing batch mean.

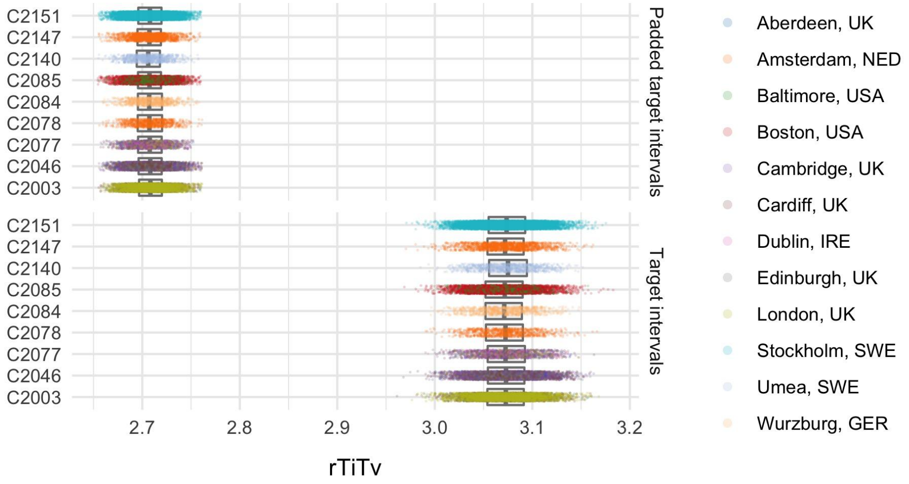


**Figure S5:** TiTv before and after further restriction to Target intervals with no padding. In each plot, jittered scatters display the distribution for each sequencing batch, coloured according to sample collection. Boxplots behind the scatter display the median and interquartile range for each sequencing batch.

| Filter | Variants | % |
|---|---|---|
| Variants with < 7 alleles | 37,344,246 | 100.0 |
| Failing VQSR | 100,742 | 0.3 |
| In LCRs | 1,215,218 | 3.3 |
| Outside padded target interval | 27,119,165 | 72.6 |
| Invariant sites after initial variant and genotype filters | 3,117,961 | 8.3 |
| Invariant sites after sample filters | 1,051,421 | 2.8 |
| Overall variant call rate < 0.97 | 737,072 | 2.0 |
| Overall variant case call rate < 0.97 | 716,709 | 1.9 |
| Overall variant control call rate < 0.97 | 743,659 | 2.0 |
| Difference between case and control variant call rate < 0.02 | 232,341 | 0.6 |
| Variants failing HWE filter | 1,083,479 | 2.9 |
| **Variants remaining after all filters** | **5,104,759** | **13.7** |

**Table S2**: Summary of variant filters. Moving down through the rows of the table, we move through QC filters described in the methods section. Full details and code are provided at astheeggeggs.github.io/BipEx.

| Filter | Samples | Bipolar cases | Controls | % |
|---|---|---|---|---|
| Initial samples in vcf | 39,618 | 16,486 | 17,212 | 100.0 |
| Unable to obtain both phenotype and sequence information | 2 | - | - | 0.0 |
| Unknown phenotype | 32 | - | - | 0.1 |
| Low coverage or high contamination | 133 | 72 | 54 | 0.3 |
| Sample call rate < 0.93 | 185 | 124 | 53 | 0.5 |
| % FREEMIX contamination > 0.02 | 268 | 146 | 104 | 0.7 |
| % chimeric reads > 0.015 | 152 | 49 | 100 | 0.4 |
| Mean DP < 30 | 20 | 5 | 12 | 0.1 |
| Mean GQ < 55 | 56 | 28 | 25 | 0.1 |
| Samples with sex swap | 238 | 147 | 52 | 0.6 |
| Related samples for removal | 1,716 | 792 | 688 | 4.3 |
| PCA based filters | 2,880 | 1,120 | 1,422 | 7.3 |
| Within batch Ti/Tv ratio outside 3 standard deviations | 100 | 50 | 42 | 0.3 |
| Within batch Het/HomVar ratio outside 3 standard deviations | 150 | 66 | 58 | 0.4 |
| Within batch Insertion/Deletion ratio outside 3 standard deviations | 93 | 31 | 48 | 0.2 |
| Within location *n* singletons outside 3 standard deviations | 443 | 151 | 236 | 1.1 |
| **Samples after final sample filters** | **33,527** | **13,933** | **14,422** | **84.6** |

**Table S3**: Summary of sample filters. Moving down through the rows of the table, we move through QC filters described in the methods section. Full details and code are provided at astheeggeggs.github.io/BipEx.

#### Final sample counts for analysis

Following data curation and quality control, the resultant composition of the samples by collection and bipolar subtype is summarised in Table S4.

| Location | BD | BD1 | BD2 | BDNOS | SAD | BD total | Controls | BD total and controls |
|---|---|---|---|---|---|---|---|---|
| Aberdeen, UK | 0 | 0 | 0 | 0 | 0 | 0 | 322 | 322 |
| Amsterdam, NED | 1 | 951 | 155 | 9 | 19 | 1,116 | 1,359 | 2,475 |
| Baltimore, USA | 3 | 254 | 6 | 4 | 0 | 267 | 41 | 308 |
| Boston, USA | 248 | 1,503 | 279 | 404 | 31 | 2,434 | 2,544 | 4,978 |
| Cambridge, UK | 0 | 0 | 0 | 0 | 0 | 0 | 2,656 | 2,656 |
| Cardiff, UK | 64 | 1,301 | 681 | 62 | 65 | 2,108 | 1,006 | 3,114 |
| Dublin, IRE | 0 | 150 | 0 | 0 | 11 | 150 | 7 | 157 |
| Edinburgh, UK | 298 | 317 | 94 | 2 | 6 | 711 | 58 | 769 |
| London, UK | 212 | 1,169 | 350 | 0 | 144 | 1,731 | 1,082 | 2,813 |
| Stockholm, SWE | 128 | 2,095 | 1,595 | 791 | 1 | 4,609 | 4,530 | 9,139 |
| Umea, SWE | 0 | 297 | 141 | 3 | 0 | 441 | 426 | 867 |
| Wurzburg, GER | 7 | 201 | 145 | 13 | 0 | 366 | 391 | 757 |
| **Total** | **961** | **8,238** | **3,446** | **1,288** | **277** | **13,933** | **14,422** | **28,355** |

**Table S4**: Detailed summary of subtype sample contributions across locations following variant and sample QC. BD=BD without a fine subclassification, BD1=bipolar I disorder, BD2=bipolar II disorder, BDNOS=bipolar disorder not otherwise specified, SAD=schizoaffective disorder, BD total = BD+BD1+BD2_BDNOS, BD total and controls=BD total+controls (excluding SAD).

### Variant annotation

We use the Ensembl Variant Effect Predictor (VEP) (*39*) version 95 with the loftee plugin to annotate variants against GRCh38 using hail, including SIFT (*40*) and Polyphen2 scores (*41*), according to the GENCODE v19 reference. The configuration file available in google cloud: gs://hail-us-vep/vep95-GRCh38-loftee-gcloud.json. In addition, we annotate with version 2.1.1 gnomAD site annotations (*42*) and MPC scores (*43*) after lifting the genome coordinates over to GRCh38. MPC is an aggregate score which uses ExAC to identify sub-genic regions that are depleted of missense variation in combination with existing metrics to create a composite predictor. Finally, we annotate with Combined Annotation Dependent Depletion (CADD) version 1.4 (*43*, *44*), and annotate constraint using the gnomAD loss of function (LOF) metrics table from release 2.1.1 (*42*). We then process the VEP annotated consequences, and define variant specific consequences and gene annotations as the most severe consequence of a canonical transcript on which that variant lies. We then assign variants (where possible) to four distinct consequence classes: protein truncating variant (PTV), missense, synonymous, and non-coding as defined in Table S5. We then subdivide missense variants into 'damaging missense' if both the polyphen prediction is 'probably damaging' and the SIFT prediction is 'deleterious', and 'other missense' otherwise.

| **Consequence class** | **VEP consequences** |
|---|---|
| PTV | Transcript ablation, splice acceptor variant, splice donor variant, stop gained, frameshift variant. |
| Missense | Stop lost, start lost, transcript amplification, inframe insertion, inframe deletion, missense variant, protein altering variant, splice region variant. |
| Synonymous | Incomplete terminal codon variant, stop retained variant, synonymous variant. |
| Non-coding | Coding sequence variant, mature miRNA variant, 5' UTR variant, 3' UTR variant, non-coding transcript exon variant, intron variant, NMD transcript variant, non-coding transcript variant, upstream gene variant, downstream gene variant , TFBS ablation, TFBS amplification, TF binding site variant, regulatory region ablation, regulatory region amplification, feature elongation, regulatory region variant, feature truncation, intergenic variant. |

**Table S5**: Consequence classes defined based VEP annotation.

### Exome-wide burden analyses

We ran a series of logistic regressions to test for an association between putatively damaging rare variation and case status, and linear regressions to test for an association between case status and excess burden of damaging variation. Note that both tests will result in near identical *P*-values; the motivation here is to ascertain two effect size parameters of rare-variant burden. We then sought to hone in on more recent mutations by restricting to rare variation not present in the non-psych portion of the gnomAD database, and perform the same collection of association tests. Furthermore, we leveraged evolutionary constraint models to enrich for deleterious variation by testing for enrichment of missense variation with MPC score ≥ 2 (representing the top ~3.9% pathogenicity of missense variation (*43*), and restricting our PTV enrichment tests within genes most likely to be loss-of-function intolerant (*p*LI ≥ 0.9).

Throughout, we test for a signal of enrichment of synonymous, and other-missense as a negative control to confirm that our burden model was well calibrated. For each collection of regressions, we include sex, ten PCs and overall burden of MAC ≤ 5 variants in the dataset following the imposed restrictions (e.g. not in gnomAD non-psych). In each case, regressions were robust to incorporation of the overall burden covariate: the overall observed patterns did not change if we controlled for overall coding burden or did not control for overall burden.

To ensure that the results in the full dataset were not driven by artefacts introduced by jointly analysing multiple cohorts or residual population structure, we also ran burden tests within each location (Table S7) and meta-analysed these results. We observed consistent results across the cohorts, and found that estimated odds ratios and excess burden between the joint analysis and meta-analysis were roughly equivalent.

#### Schizophrenia as a positive control for damaging rare burden analysis

In the case of schizophrenia, multiple studies have shown enrichment of rare damaging coding variation in cases over controls (*45*, *46*). As a positive control, we considered the subset of schizophrenia cases in the BipEx cohort and tested for enrichment of putatively damaging variation in these loss of function intolerant (*p*LI > 0.9) genes and replicated this result (OR = 1.28, $p = 1.9 \times 10^{-10}$).

### Age of onset definitions

Three definitions for age of onset were available for subsets of the data and considered for analysis: age at first symptoms, age at first diagnosis, and age at first impairment. In each case, two distinct age encodings were used:

1. < 18; 18-40; 40+.
2. < 12; 12-24; 24+.

**Cardiff, UK**
Age at first symptoms: SCAN (*3*) interview and case records; age of first clinically significant symptoms due to affective/psychotic illness was used to define encodings 1 and 2.
Age at first impairment: SCAN (*3*) interview and case records; age of first clinically significant impairment due to affective/psychotic illness was used to define encodings 1 and 2.

**Boston, USA**
Age of diagnosis: A regular expression algorithm extracting mention by clinician in an inpatient or outpatient note (*14*, *47*). Age of onset must be explicitly mentioned by a physician in a clinical note. Results were used to define encoding 1.

**London, UK**
Age of first impairment. OPCRIT (*3*, *4*) question 4 of the DPIM BPAD questionnaire (github.com/astheeggeggs/BipEx/DPIM_BPAD.docx): age of onset, defined as the earliest age at which medical advice was sought for psychiatric reasons or at which symptoms began to cause subjective distress or impair functioning, provided to the nearest year. Age was used to define encodings 1 and 2.

**Stockholm, SWE**
**SWEBIC (Swedish Bipolar Cohort Collection), SWE**
**SBP**
Age at first symptoms: Age at first sign of psychiatric disorder as recorded in the ADE
Age at first diagnosis: Age at first contact with healthcare professionals for mental health issues as recorded in ADE.
**BipoläR**:
Age at first symptoms: Age at first signs of mental health problems or psychiatric disorder as recorded in the QA-register stratified by < 8 years of age, 8-11 yrs, 12-17 yrs, 18-24 yrs, > 24 years of age;
Age at first diagnosis: Question at telephone interview: “How old were you at your first contact with health care professionals due to mental health issues / a psychiatric disorder?”
**HDR**:
Age at first symptoms: Age at first signs of mental health problems / psychiatric disorder as recorded in the telephone interview stratified by < 8 years of age, 8-11 yrs, 12-17 yrs, 18-24 yrs, > 24 years of age.

Age at first diagnosis: Question at telephone interview: “How old were you at your first contact with health care professionals due to mental health issues / a psychiatric disorder?”

Age at first diagnosis exists as actual age but was divided according to encodings 1 and 2.
Age at first symptoms is provided according to encoding 2.

Data on age of first impairment was not collected in the Swedish cohort collection. However, a subset of Swedish data (The St. Göran Project, **SBP**) contains information on, ‘age at first health care contact for any psychiatric problem’, which herein was considered to indicate age of first impairment, and was divided according to encodings 1 and 2.

### Testing for relationship between age of onset and rare variant burden

To test for an association between age of onset (see ‘age of onset definitions’ for full details) and burden of rare damaging variation, we first restricted our attention to the class of variation with the strongest signal for excess in cases over controls, PTVs. We considered only ‘age at first impairment’ (Table S10) as this was the definition with the largest amount of available data: 3,677 in both encoding 1 and 2. Using these two encodings, we further split the age of first impairment categories into five discrete bins: < 12, 12-18, 18-24, 24-40 and > 40. We tested all 10 possible ‘younger bin’ vs ‘older bin’ pairs across this partition to check for differences in MAC ≤ 5 PTV burden, MAC ≤ 5 not in gnomAD PTV burden, and MAC ≤ 5 not in gnomAD PTV in *p*LI ≥ 0.9 burden, using Kolmogorov-Smirnov tests. We also used Fisher’s exact tests to test for an association between carrier status for the damaging rare PTV categories between the ‘younger’ and ‘older’ bins.

| Location | Age First Impairment <12 | Age First Impairment 12-24 | Age First Impairment >24 | Total | Age First Impairment <18 | Age First Impairment 18-40 | Age First Impairment >40 | Total |
|---|---|---|---|---|---|---|---|---|
| Cardiff, UK | 80 | 824 | 404 | 1,308 | 469 | 782 | 57 | 1,308 |
| London, UK | 78 | 978 | 752 | 1,808 | 446 | 1,188 | 174 | 1,808 |
| Stockholm, SWE | 26 | 256 | 279 | 561 | 135 | 355 | 71 | 561 |
| **Total** | **184** | **2,058** | **1,435** | **3,677** | **1,050** | **2,325** | **302** | **3,677** |

**Table S6**: Age of onset ‘age of first impairment’ data. We have data split according to two encodings as described in ‘age on onset definitions’ in three of the BipEx cohorts.

### Psychosis definitions

Psychosis was defined by a lifetime history of hallucinations or delusions. Presence of psychosis was evaluated differently across cohorts based on available data.

**Boston, USA**
Validated Natural language processing based algorithm run on clinical notes (*14*, *47*).

**Cardiff, UK**
SCAN interview (*3*) and case records. Definite evidence of lifetime presence of psychotic symptoms and lifetime presence of individual OPCRIT (*3*, *4*) psychotic symptoms.

**London, UK**
OPCRIT (*3*, *4*) interview: lifetime presence of psychotic symptoms as defined by questions 52, 54, 55, 57-77 of the OPCRIT checklist detailed in the DNA polymorphisms in mental illness (DPIM) bipolar affective disorder (BPAD) questionnaire (github.com/astheeggeggs/BipEx/DPIM_BPAD.docx).

**Stockholm, SWE**
**SWEBIC (Swedish Bipolar Cohort Collection), SWE**
**SBP:** ADE question: any psychotic disorder?
**BipoläR and HDR:** During a structured telephone interview that research nurses conducted, "have you ever lost touch with reality (i.e. have heard or seen things that others have not seen) or experienced things that you later realized were not real?" was asked. Patients were defined as having psychosis if the answer to this question was clear-cut 'yes', and not having psychosis if doubtful.

**Before restricting to samples with high quality sequence data**

| | Psychosis | | | | | No psychosis | | | | | |
|---|---|---|---|---|---|---|---|---|---|---|---|
| | Bipolar Disorder | | | | | Bipolar Disorder | | | | | |
| **Location** | **BD** | **BD1** | **BD2** | **BDNOS** | **SAD** | **BD** | **BD1** | **BD2** | **BDNOS** | **SAD** | **BD total** |
| Boston, USA | 13 | 438 | 82 | 100 | 34 | 6 | 105 | 27 | 34 | 0 | 805 |
| Cardiff, UK | 40 | 994 | 74 | 6 | 54 | 12 | 247 | 488 | 30 | 0 | 1,891 |
| London, UK | 12 | 869 | 86 | 0 | 128 | 6 | 343 | 226 | 0 | 9 | 1,542 |
| Stockholm, SWE | 0 | 1,349 | 497 | 315 | 0 | 0 | 742 | 1,142 | 510 | 0 | 4,555 |
| Wurzburg, GER | 0 | 47 | 11 | 1 | 0 | 7 | 169 | 148 | 14 | 0 | 397 |
| **Total** | **65** | **3,697** | **750** | **422** | **216** | **31** | **1,606** | **2,031** | **588** | **9** | **9,190** |

**Following restriction to samples with high quality sequence data**

| | Psychosis | | | | | No psychosis | | | | | |
|---|---|---|---|---|---|---|---|---|---|---|---|
| | Bipolar Disorder | | | | | Bipolar Disorder | | | | | |
| **Location** | **BD** | **BD1** | **BD2** | **BDNOS** | **SAD** | **BD** | **BD1** | **BD2** | **BDNOS** | **SAD** | **BD total** |
| Boston, USA | 9 | 303 | 54 | 66 | 22 | 6 | 74 | 19 | 23 | 0 | 554 |
| Cardiff, UK | 29 | 842 | 65 | 6 | 51 | 11 | 216 | 438 | 28 | 0 | 1,635 |
| London, UK | 11 | 770 | 79 | 0 | 118 | 6 | 317 | 219 | 0 | 9 | 1,402 |
| Stockholm, SWE | 0 | 1,193 | 453 | 279 | 0 | 0 | 659 | 1,034 | 442 | 0 | 4,060 |
| Wurzburg, GER | 0 | 44 | 10 | 1 | 0 | 7 | 157 | 135 | 12 | 0 | 366 |
| **Total** | **49** | **3,152** | **661** | **352** | **191** | **30** | **1,423** | **1,845** | **505** | **9** | **8,017** |

**Table S7**: Breakdown of psychosis diagnosis information across BipEx cohorts available in the phenotype data, and following destruction to the analysis ready dataset. BD=BD without a fine subclassification, BD1=bipolar I disorder, BD2=bipolar II disorder, BDNOS=bipolar disorder not otherwise specified, SAD=schizoaffective disorder, BD+SAD=bipolar disorder and schizoaffective disorder combined, SCZ=schizophrenia, other=other unspecified case, unknown=unknown case status.

### Gene-set enrichment analysis

To test for enrichment of damaging rare variation in gene sets, we regressed case status against MAC ≤ 5 burden of each variant class (Table S5) in each gene set of interest using logistic regression, and regressed burden of each variant class on case-status using linear regression. We correct for overall MAC ≤ 5 coding burden in the gene set, sex, and first ten PCs. We also included overall rare coding burden as a covariate ensured that any signal was significant above overall rare coding differences between cases and controls in the analysed gene set. All cohorts were analysed together.

Following the observation of enrichment of brain expressed genes in the initial gene-set analysis, we sought to refine the signal. In the collection of GTEx tissue specific gene-sets defined by (*48*), a subset of 13 are brain regions (italicised in Table S8). We tested for enrichment of MAC ≤ 5 PTVs in these GTEx gene-sets defined as having the strongest tissue specific expression using logistic regression, again controlling for 10 PCs, sex and MAC ≤ 5 coding burden. The 43 tested genesets are available for download at https://data.broadinstitute.org/alkesgroup/LDSCORE/LDSC_SEG_ldscores/. Results are displayed in Figure 2A.

| Tissue | Tissue category | Broad tissue category | Number of samples |
|---|---|---|---|
| Bladder | Bladder | Other | 11 |
| Kidney Cortex | Kidney | Other | 32 |
| Minor Salivary Gland | Salivary Gland | Other | 57 |
| *Brain Substantia nigra* | *Brain* | CNS | 63 |
| *Brain Spinal cord (cervical c-1)* | *Brain* | CNS | 70 |
| *Brain Amygdala* | *Brain* | CNS | 72 |
| *Brain Anterior cingulate cortex (BA24)* | *Brain* | CNS | 84 |
| Small Intestine Terminal Ileum | Small Intestine | Digestive | 88 |
| *Brain Hippocampus* | *Brain* | CNS | 94 |
| *Brain Hypothalamus* | *Brain* | CNS | 96 |
| *Brain Putamen (basal ganglia)* | *Brain* | CNS | 97 |
| Pituitary | Pituitary | Endocrine | 103 |
| Spleen | Spleen | Blood/Immune | 104 |
| *Brain Cerebellar Hemisphere* | *Brain* | CNS | 105 |
| *Brain Frontal Cortex (BA9)* | *Brain* | CNS | 108 |
| *Brain Nucleus accumbens (basal ganglia)* | *Brain* | CNS | 113 |
| *Brain Cortex* | *Brain* | CNS | 114 |
| *Brain Caudate (basal ganglia)* | *Brain* | CNS | 115 |
| Liver | Liver | Liver | 119 |
| *Brain Cerebellum* | *Brain* | CNS | 125 |
| Artery Coronary | Blood Vessel | Cardiovascular | 133 |
| Adrenal Gland | Adrenal Gland | Endocrine | 145 |
| Colon Sigmoid | Colon | Digestive | 149 |
| Esophagus Gastroesophageal Junction | Esophagus | Digestive | 153 |
| Pancreas | Pancreas | Other | 171 |
| Stomach | Stomach | Digestive | 193 |
| Heart Atrial Appendage | Heart | Cardiovascular | 194 |
| Colon Transverse | Colon | Digestive | 196 |
| Breast Mammary Tissue | Breast | Other | 214 |
| Heart Left Ventricle | Heart | Cardiovascular | 218 |
| Artery Aorta | Blood Vessel | Cardiovascular | 224 |
| Adipose Visceral (Omentum) | Adipose Tissue | Adipose | 227 |
| Esophagus Muscularis | Esophagus | Digestive | 247 |
| Skin Not Sun Exposed (Suprapubic) | Skin | Other | 250 |
| Esophagus Mucosa | Esophagus | Digestive | 286 |
| Nerve Tibial | Nerve | Other | 304 |
| Lung | Lung | Other | 319 |
| Thyroid | Thyroid | Endocrine | 322 |
| Artery Tibial | Blood Vessel | Cardiovascular | 332 |
| Adipose Subcutaneous | Adipose Tissue | Adipose | 350 |
| Skin Sun Exposed (Lower leg) | Skin | Other | 357 |
| Whole Blood | Blood | Blood/Immune | 393 |
| Muscle Skeletal | Muscle | Musculoskeletal/connective | 430 |

**Table S8:** GTEx tissue information for analysed (*48*) GTEx gene-sets. The 43 tested genesets are available at data.broadinstitute.org/alkesgroup/LDSCORE/LDSC_SEG_ldscores/.

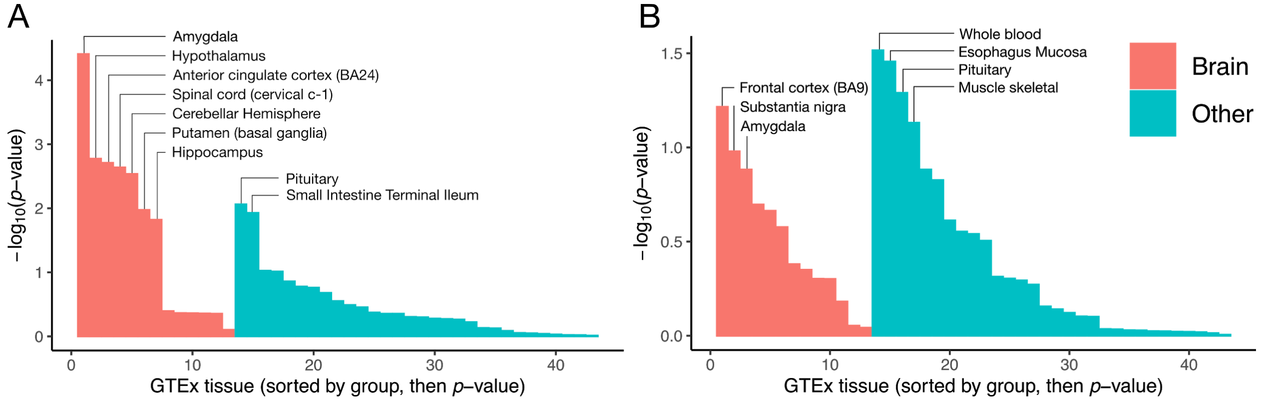


**Figure S6:** Enrichment of ultra-rare PTVs in BD1 and BD2 cases over controls in tissue-specific expression genesets. Gene-sets are defined in (*48*) in detail. Bars are ordered first by whether they are a brain-tissue, and then by *P*-value. A. displays the results for BD1, B. displays the results for BD2.

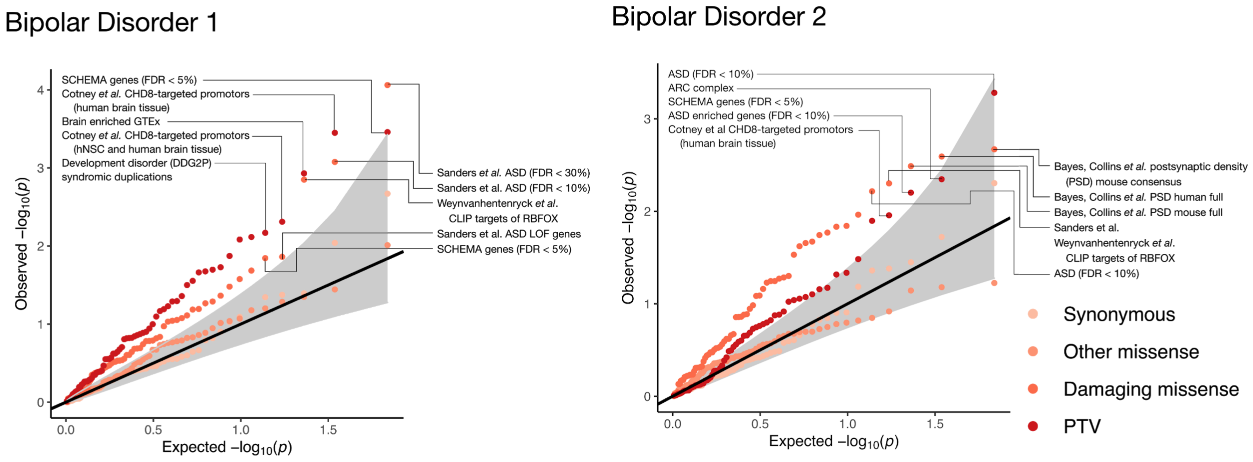


**Figure S7:** Enrichment of ultra-rare variants in targeted 68 gene-sets taken from the literature. The left plot shows enrichment in BD1, the right plot shows enrichment in BD2. Top PTV and damaging missense gene-sets are labelled. Classes of variants tested in each gene-set are coloured according to the legend.

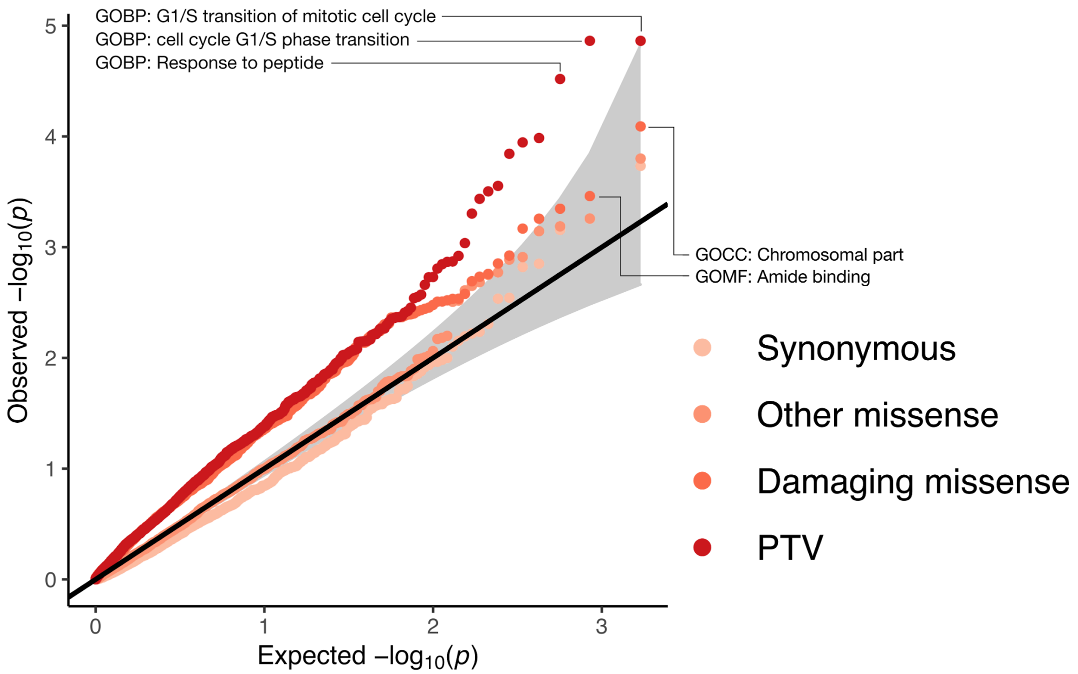


**Figure S8:** QQ plot of *P*-values testing for enrichment of ultra-rare variants in 1,697 gene-sets taken from derived from large pathway databases including Gene Ontology (GO), REACTOME and KEGG). Top PTV and damaging missense gene-sets are labelled. Classes of variants tested in each gene-set are coloured according to the legend.

### Gene-based analysis approach

In order to increase power for gene discovery, we filter down to variants not present in the non-psych portion of the gnomAD dataset (*42*), and we further enriched for pathogenic variants by restricting our analysis to variants with MAC ≤ 5. We then examine case-control enrichment of PTVs (transcript ablation, splice acceptor variant, splice donor variant, stop gained and frameshift variant; Table S5) or damaging missense variants (missense variants (Table S5) annotated as 'probably damaging' in PolyPhen and 'deleterious' in SIFT). We further restricted our analysis to the coding exons within the target intervals of the illumina capture, to reduce potential for artefacts which could potentially be induced due to differential coverage across batches in any padded target interval, using synonymous, and other missense ultra-rare variants in each gene as the negative control (Figure S11-12).

Throughout, we use Fisher's exact tests in each gene. We considered a Cochran–Mantel–Haenszel (CMH) test, using the strata defined by broad geographic location. We use a permutation approach to determine the null distribution of test statistics throughout our gene based analysis, and evaluate QQ plots of synonymous and other-missense ultra-rare variants to ensure that tests are well-calibrated (Figure S11-12). We used Fisher's exact tests in our primary analysis, as tests showed the strongest power and also had well calibrated QQ plots across annotation categories (Figure S11-15).

#### CMH and Fisher's exact test for gene based tests

We tested for an excess of ultra-rare variation (MAC ≤ 5 and not present in the non-psych portion of the gnomAD dataset) in each gene using both Fisher's exact and Cochran-Mantel-Haenszel (CMH) tests for each phenotype. Given that we did not observe excess burden in missense variants with high MPC (>3 or >2) in bipolar cases over controls exome wide (in contrast to schizophrenia; (*49*)), we did not test a weighted summation of counts across consequence categories. For each gene, each sample was assessed for carrier status for each of the following consequence classes: synonymous, other missense, damaging missense, and PTV (Table S5); individuals harbouring at least one copy in the consequence class under analysis were counted as carriers. These counts were then taken through to define 2 × 2 and 2 × 2 × 6 contingency tables for Fisher's exact and CMH tests respectively, using location as strata, see below. To ensure that our tests were well calibrated, we randomly permuted case labels (within stratum for CMH) for each gene and reran the test 20 times across all genes and keep track of the summation of the ordered vectors of *P*-values up to that permutation, before taking an average at the last permutation. This vector of length |*n* genes| then defines our expected distribution of *P*-values. Fisher's exact test *P*-values and odds-ratio for carrier status are displayed in the gene results tables on the browser: bipex.broadinstitute.org.

| Location stratum | Cohort |
|---|---|
| UK/Ireland | Aberdeen, UK<br>Cambridge, UK<br>Cardiff, UK<br>Dublin, IRE<br>Edinburgh, UK<br>London, UK |
| Germany | Wurzburg, GER |
| USA | Baltimore, USA<br>Boston, USA |
| Netherlands | Amsterdam, NED |
| Sweden, Stockholm | Stockholm, SWE |
| Sweden, Umea | Umea, SWE |

#### Robustness of gene-based analysis

To ensure that our tests were robust, we performed a series of checks to see if the Fisher's exact (Figures S11-12), and Cochran-Mantel–Haenszel (CMH) tests results showed an elevated false positive rate. In both tests, we observed the expected null *P*-value distribution in the collection of gene-based tests when analysing synonymous and 'other-missense' variants with MAC ≤ 5 not in gnomAD non-psych. To further test calibration of the test statistic, we filtered to genes where we are well powered to detect differences between BD cases and controls. We examined case-control enrichment of synonymous ultra-rare variants in genes with an allele count of > 20 and > 50 and compared observed *P*-value to the uniform expectation (Figure S15). In each, we did not observe inflation of the test statistic.

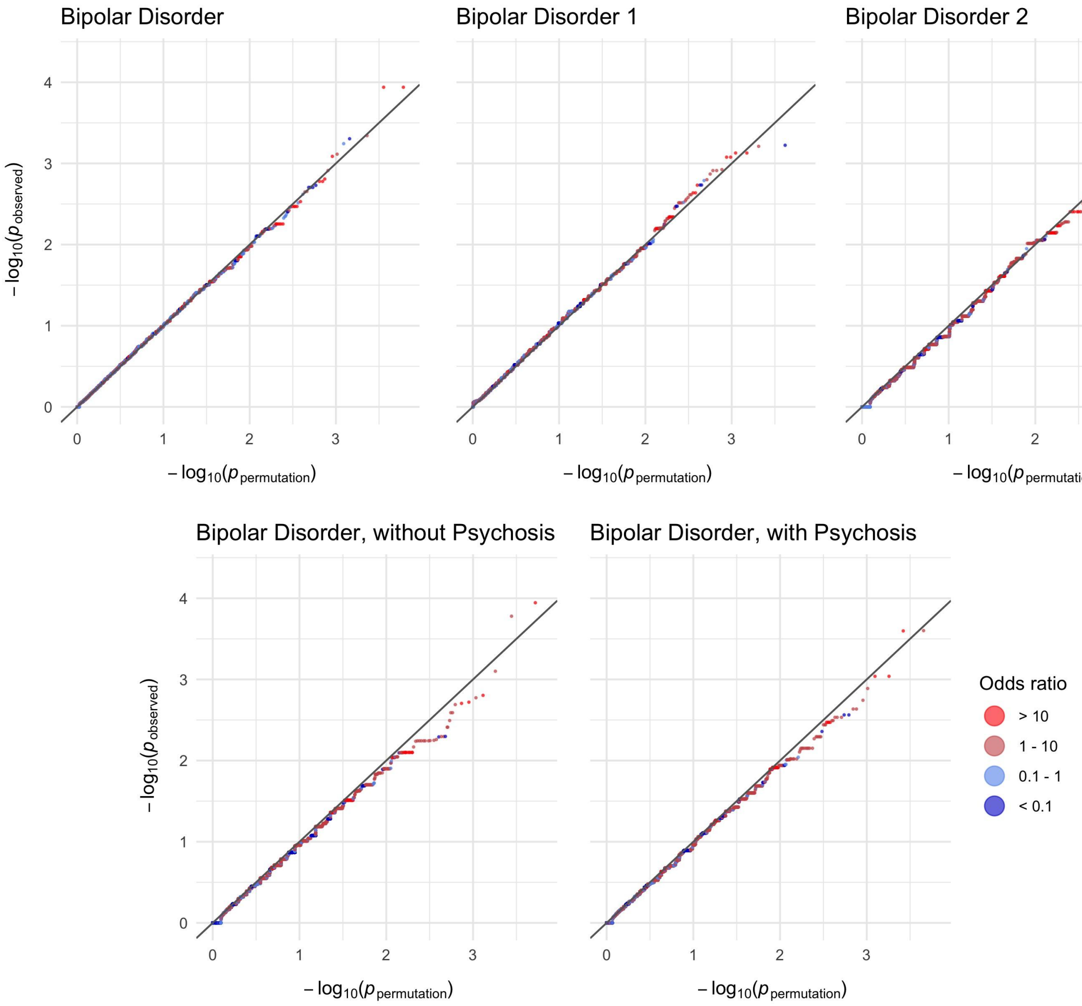


**Figure S9:** MAC ≤ 5 not in gnomAD non-psych synonymous variants in 13,933 cases and 14,422 controls: QQ plots. Observed -log10 *p*-values are plotted against permutation *p*-values according to the procedure described in the methods; gene-based analysis. Points are coloured according to the discrete scale displayed in the legend.

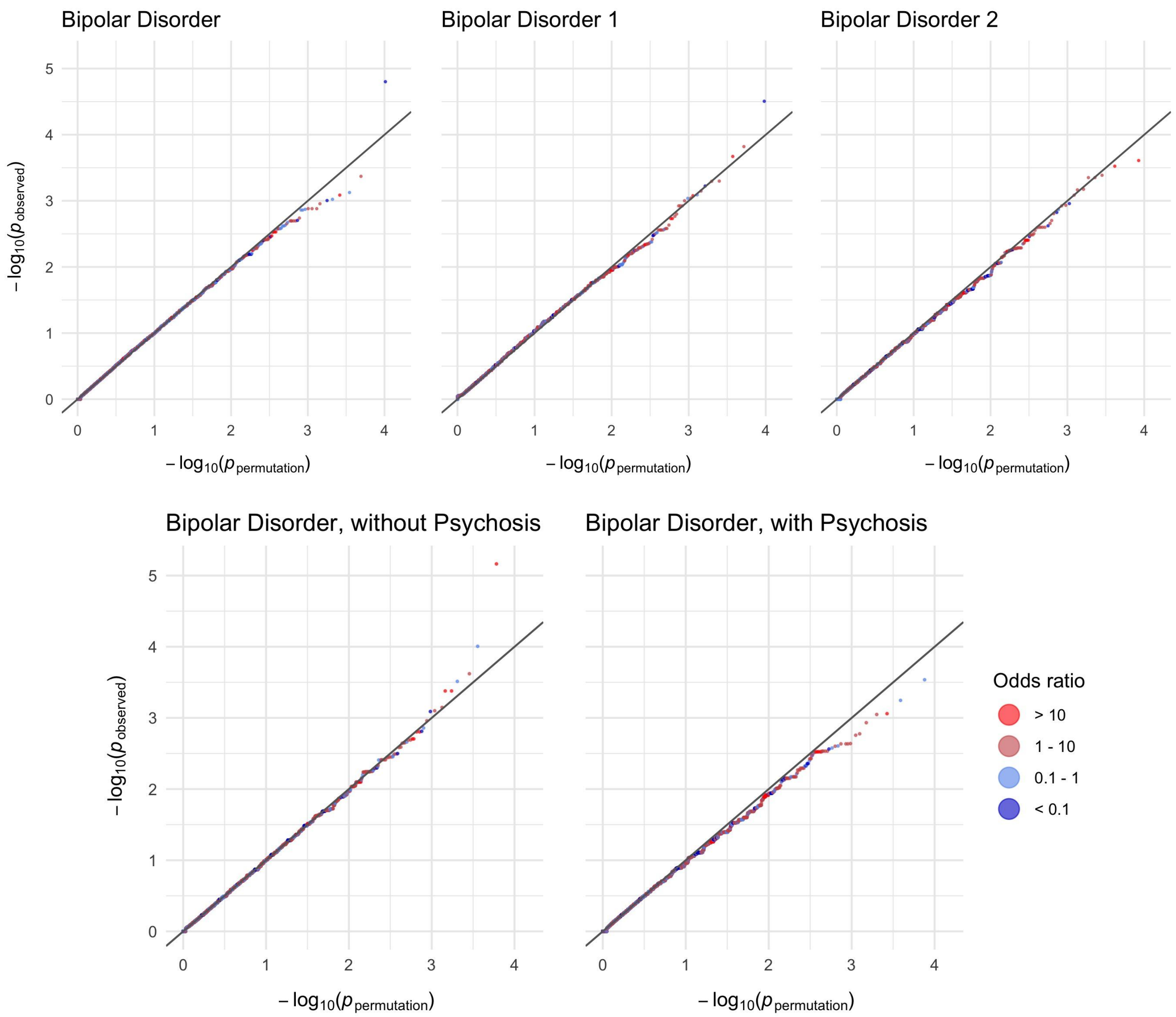


**Figure S10:** MAC ≤ 5 not in gnomAD non-psych other missense variants in 13,933 cases and 14,422 controls: QQ plots. Observed -log10 *p*-values are plotted against permutation *p*-values according to the procedure described in the methods; gene-based analysis. Points are coloured according to the discrete scale displayed in the legend.

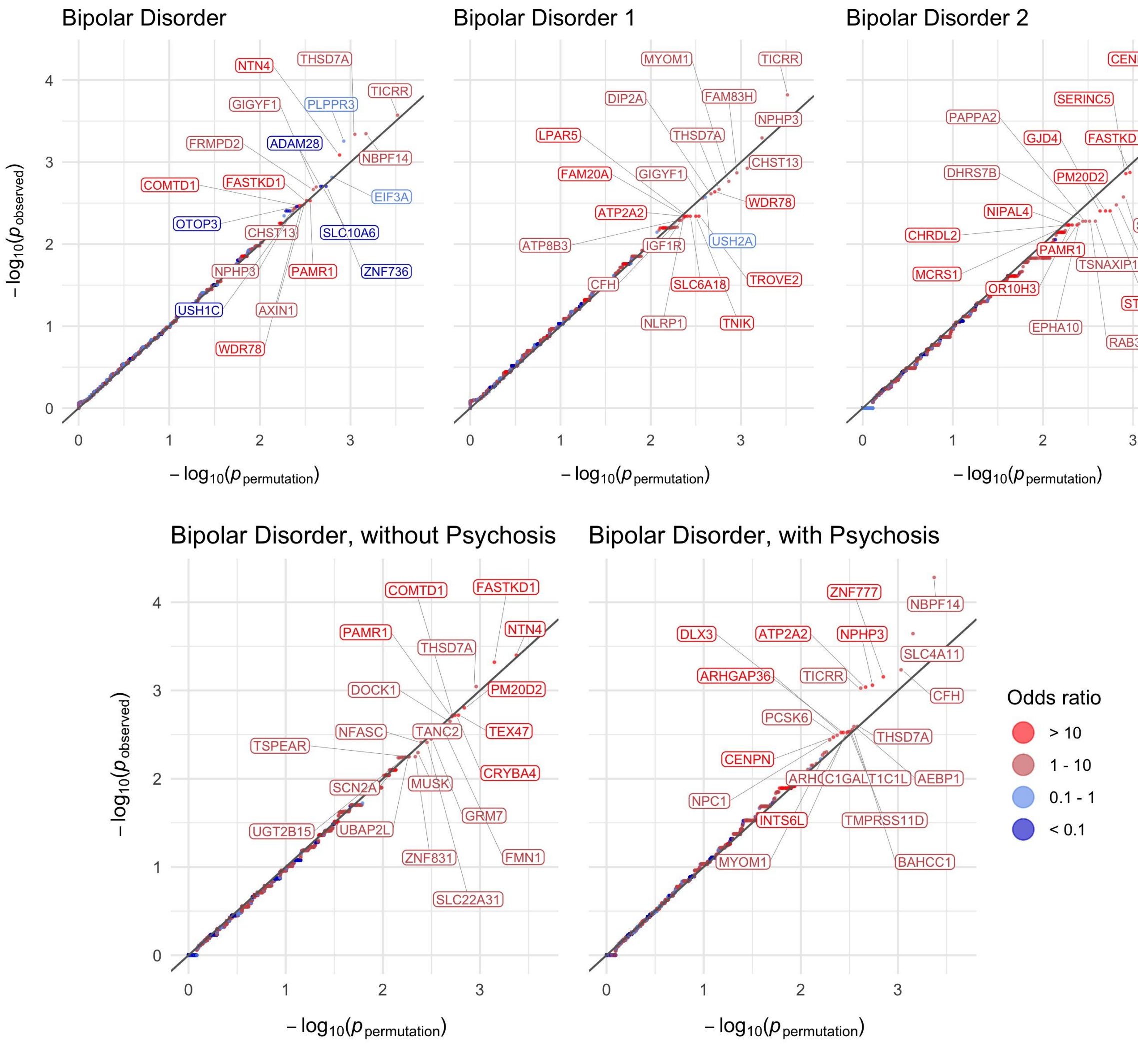


**Figure S11:** MAC ≤ 5 not in gnomAD non-psych damaging missense variants in 13,933 cases and 14,422 controls: QQ plots. Observed -log10 *P*-values are plotted against permutation *P*-values according to the procedure described in the methods; gene-based analysis. Points are coloured according to the discrete scale displayed in the legend. In each panel, the gene symbols of the top 20 genes by *P*-value are labelled.

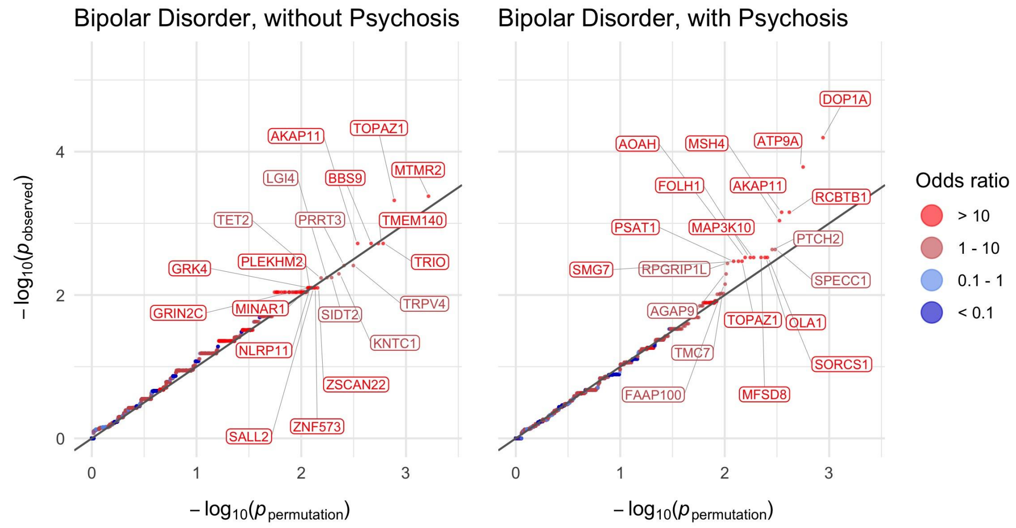


**Figure S12:** MAC ≤ 5 not in gnomAD non-psych PTVs in 13,933 cases and 14,422 controls: QQ plots. Observed -log10 *P*-values are plotted against permutation *P*-values according to the procedure described in the methods; gene-based analysis. Points are coloured according to the discrete scale displayed in the legend. In each panel, the gene symbols of the top 20 genes by *P*-value are labelled.

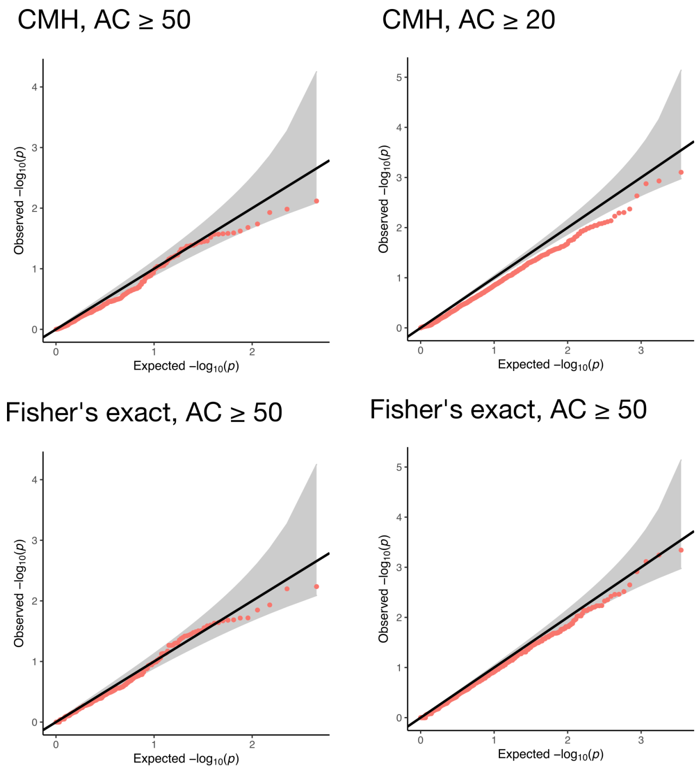


**Figure S13**: MAC ≤ 5 not in gnomAD non-psych synonymous variants in 13,933 cases and 14,422 controls: QQ plots for BD in genes with ultra-rare synonymous counts about 20 and 50 across BD cases and controls. Observed -log10 *P*-values are plotted against expected *P*-values using a uniform distribution. The first and second rows show *P*-values obtained via a CMH and Fisher's exact test respectively. The first a second columns restrict to genes with at least 20 and at least 50 individuals across cases and controls harbouring an ultra-rare PTV respectively.

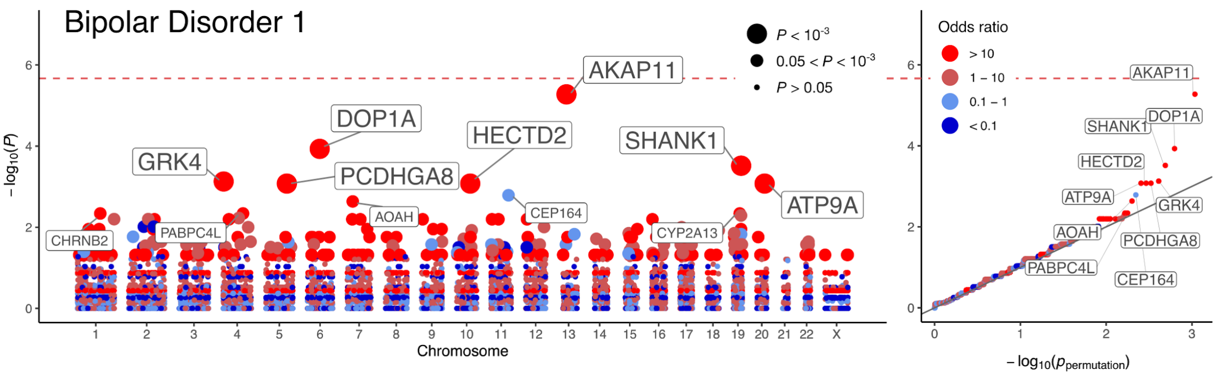


**Figure S14: Results of the analysis of ultra-rare PTVs in BD1: 8,238 cases and 14,422 controls.** Gene based Manhattan and associated QQ plot for BD1. -$\log_{10}$ *P*-values obtained via Fisher's exact tests are plotted against genetic position for each of the analysed genes. In the QQ plots, observed -$\log_{10}$ *P*-values are plotted against permutation *P*-values according to the procedure described in the supplementary materials: gene-based analysis approach. Points are coloured according to the discrete scale displayed in the legend. In the Manhattan plot and QQ plot, the gene symbols of top genes by *P*-value are labelled. Points in the Manhattan plot are sized according to *P*-value as displayed in the legend.

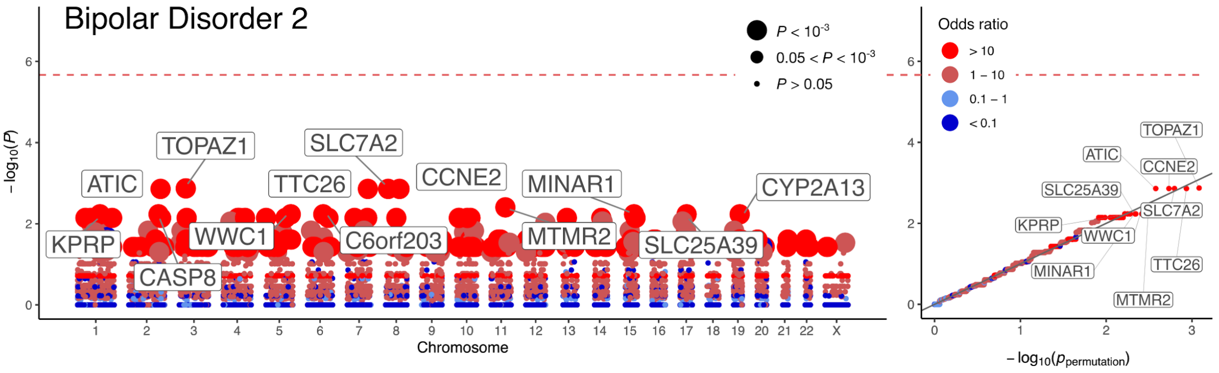


**Figure S15: Results of the analysis of ultra-rare PTVs in BD2: 3,446 cases and 14,422 controls.** Gene based Manhattan and associated QQ plot for BD1. -$\log_{10}$ *P*-values obtained via Fisher's exact tests are plotted against genetic position for each of the analysed genes. In the QQ plots, observed -$\log_{10}$ *P*-values are plotted against permutation *P*-values according to the procedure described in the supplementary materials: gene-based analysis approach. Points are coloured according to the discrete scale displayed in the legend. In the Manhattan plot and QQ plot, the gene symbols of top genes by *P*-value are labelled. Points in the Manhattan plot are sized according to *P*-value as displayed in the legend.

### Combining SCHEMA and BipEx data in meta-analysis

To examine the extent of shared ultra-rare PTV signal between BD and SCZ we ran separate Fisher and CMH tests for BipEx and SCHEMA separately and meta-analysed the results using weighted *Z*-scores, weighing by effective sample sizes. Fisher's exact and CMH two-sided *P*-values were halved and converted to signed *Z*-scores using the OR to define sign. Weighted *Z*-score were then evaluated:

$$Z = \frac{\sum_{i=1}^{m} w_i Z_i}{\sqrt{\sum_{i=1}^{m} w_i^2}},$$

where $w_i = \sqrt{N_{eff,\,i}}$, $N_{eff,\,i} = 4Np_{case,\,i}(1 - p_{case,i})$, and $p_{case,\,i}$ is the case proportion in the $i^{th}$ cohort. Associated *P*-values were then evaluated. As the UK and Ireland controls were present as controls for the SCHEMA study, these controls were excluded from the analysis.

| | BD (BipEx) | | | | SCZ (SCHEMA) | | | | Combined | |
|---|---|---|---|---|---|---|---|---|---|---|
| **Gene** | **Case count BD/BD1/BD2** BD $n$ = 13,933 BD1 $n$ = 8,238 BD2 $n$ = 3,446 | **Control count** $n$ = 14,422 | ***P*-value** | **OR** | **Case count** $n$ = 24,248 | **Control count** $n$ = 91,960 | ***P*-value** | **OR** | **OR** | **Meta *P*-value** |
| *AKAP11* | 16/12/2 | 0 | $1.15 \times 10^{-5}$ | ∞ | 17 | 13 | $2.02 \times 10^{-5}$ | 5.60 | 7.06 | $2.83 \times 10^{-9}$ |
| *DOP1A* | 15/11/2 | 1 | $2.22 \times 10^{-4}$ | 15.54 | 19 | 43 | $1.47 \times 10^{-1}$ | 1.59 | 2.11 | $1.44 \times 10^{-4}$ |
| *PCDHGA8* | 11/7/1 | 0 | $4.02 \times 10^{-4}$ | ∞ | 6 | 44 | $2.19 \times 10^{-1}$ | 0.54 | 0.99 | $3.38 \times 10^{-3}$ |
| *SHANK1* | 10/8/1 | 0 | $8.19 \times 10^{-4}$ | ∞ | 4 | 4 | $4.43 \times 10^{-1}$ | 2.90 | 6.99 | $9.71 \times 10^{-3}$ |
| *TOPAZ1* | 12/6/5 | 1 | $1.56 \times 10^{-3}$ | 12.43 | 2 | 3 | $6.67 \times 10^{-1}$ | 0.93 | 3.93 | $2.51 \times 10^{-3}$ |
| *ATP9A* | 9/7/2 | 0 | $1.66 \times 10^{-3}$ | ∞ | 15 | 11 | $6.96 \times 10^{-4}$ | 4.08 | 5.46 | $5.36 \times 10^{-6}$ |
| *FREM2* | 4/3/1 | 19 | $2.67 \times 10^{-3}$ | 0.22 | 22 | 92 | $5.48 \times 10^{-1}$ | 0.83 | 0.65 | $3.80 \times 10^{-2}$ |
| *CHD1L* | 11/6/2 | 1 | $2.95 \times 10^{-3}$ | 11.39 | 16 | 73 | $5.99 \times 10^{-1}$ | 0.82 | 1.01 | $4.57 \times 10^{-2}$ |
| *CHRNB2* | 11/7/1 | 1 | $2.95 \times 10^{-3}$ | 11.39 | 2 | 17 | $5.54 \times 10^{-1}$ | 0.52 | 1.88 | $3.04 \times 10^{-2}$ |
| *CYP2A13* | 11/7/4 | 1 | $2.95 \times 10^{-3}$ | 11.39 | 13 | 28 | $6.30 \times 10^{-1}$ | 1.29 | 2.27 | $4.61 \times 10^{-2}$ |

**Table S9**: BipEx and SCHEMA case-control counts of the top ten most significant genes in the BipEx BD main gene-based analysis. Case and control columns denote the count of ultra-rare PTVs in the gene in the respective dataset. *P*-values are determined using Fisher's exact and CMH tests for BipEx and SCHEMA (supplementary materials: gene-based analysis approach) respectively, and meta-analysed with equal weights using Stouffer's method. BipEx: BD case count 13,933, control count 14,422. SCHEMA: schizophrenia case count 24,248, control count 91,960. The SCHEMA OR is the estimated OR averaged over strata, whereas the combined OR is the simple OR calculated by combining the BipEx and SCHEMA cases and controls.

### Lithium response

**Stockholm, SWE**

**SWEBIC (Swedish Bipolar Cohort Collection), SWE**

**SBP:** Not available.

**BipoläR and HDR:** During a structured telephone interview that research nurses conducted, patients who had been on lithium for at least 12 months were asked the following question: "What do you think of the effect (of lithium)? Do not consider side effects." Patients were partitioned according to the following response options.

0: Non-responder 'None or very doubtful effect'.
1: Partial-responder 'Doubtless effect of treatment but additional temporary or continuous treatment needed'.
2: Good-responder 'Complete response, recovered'.

**Cardiff, UK**

0. No evidence of response.
1. Subjective good response - upon interview, patients reported that lithium helped stabilise their moods.
2. Objective evidence for beneficial response, i.e., clear reduction in number and/or severity of episodes following introduction of lithium prophylaxis. (Can only be rated if at least 3 episodes of illness have occurred before lithium prophylaxis and lithium response has been observed for at least 3 years).
3. Objective evidence for excellent response to lithium prophylaxis, i.e., frequency of episodes reduced to < 10% of frequency after lithium prophylaxis and/or 2 or more episodes of illness occurring within weeks of cessation of lithium. (Can only be rated if at least 3 episodes of illness have occurred before lithium prophylaxis and lithium response has been observed for at least 5 years).

### External validation with the BSC exome data

To externally check our gene-based PTV results, we obtained PTV counts from the Bipolar sequencing consortium (BSC) (http://metamoodics.org/bsc/consortium/). Specifically, rare variant counts within the top ten genes defined by *P*-value in the Fisher's exact tests of enrichment of ultra-rare PTVs in the data were provided by the BSC. To harmonise the BSC data with BipEx, we used annotation definitions defined in Table S5. We then generate MAC ≤ 5 counts for each gene in the BSC data. Full details of the exome or whole genome sequencing platform for each cohort is summarised in Table S10. The addition of the BSC data set has some limitations. Primarily, frameshift indels were not called for a subset of the cohorts, reducing power to detect an association. Among the BSC cohorts that called indels, only the Rarebliss dataset provided indel calls. Furthermore, library preparation, sequencing platform, and variant calling differed across the BSC cohorts.

| **Study** | **Ethnicity** | **Sequencing platform** | **Library Preparation** | **Variant calling** | **BD** | **Controls** | **Total** |
|---|---|---|---|---|---|---|---|
| BRIDGES | US-Caucasian | HiSeq 2500 (WGS) | - | GotCloud analysis pipeline (*39*, *50*) | 1,712 | 1,844 | 3,556 |
| RareBLISS | US-Caucasian | HiSeq 2000/2500 | Nimblegen SeqCap EZ Exome | GATK (*32*) | 961 | 1,039 | 2,000 |
| Sweden | Swedish-Caucasian | HiSeq 2000/2500 | Agilent SureSelect Human All Exon v2 | GATK (*32*) | 831 | 1,956 | 2,787 |
| KPNC-EUR | US-Caucasian | HiSeq 2000/2500 | Nimblegen SeqCap EZ Exome | GATK (*32*) | 192 | 192 | 384 |
| KPNC-AFR | US-African American | HiSeq 2000/2500 | Nimblegen SeqCap EZ Exome | GATK (*32*) | 96 | 95 | 191 |
| KPNC-LAT | US-Latino | HiSeq 2000/2500 | Nimblegen SeqCap EZ Exome | GATK (*32*) | 98 | 100 | 198 |
| KPNC-EAS | US-East Asian | HiSeq 2000/2500 | Nimblegen SeqCap EZ Exome | GATK (*32*) | 97 | 96 | 193 |
| **Total** | | | | | **3,987** | **5,322** | **9,309** |

**Table S10:** Summary of BSC sample data.

| | BipEx | | | | BSC | |
|---|---|---|---|---|---|---|
| **Gene** | **Case count BD/BD1/BD2** BD *n*= 13,933 BD1 *n* = 8,238 BD2 *n* = 3,446 | **Control count** *n*=14,422 | ***P*-value** | **OR** | **Case count** *n*=3,987 | **Control count** *n*=5,322 |
| AKAP11 | 16/12/2 | 0 | $1.15 \times 10^{-5}$ | ∞ | 1 | 0 |
| DOP1A | 15/11/2 | 1 | $2.22 \times 10^{-4}$ | 15.54 | 0 | 1 |
| PCDHGA8 | 11/7/1 | 0 | $4.02 \times 10^{-4}$ | ∞ | 3 | 6 |
| SHANK1 | 10/8/1 | 0 | $8.19 \times 10^{-4}$ | ∞ | 1 | 0 |
| TOPAZ1 | 12/6/5 | 1 | $1.56 \times 10^{-3}$ | 12.43 | 1 | 0 |
| ATP9A | 9/7/2 | 0 | $1.66 \times 10^{-3}$ | ∞ | 2 | 1 |
| FREM2 | 4/3/1 | 19 | $2.67 \times 10^{-3}$ | 0.22 | 3 | 3 |
| CHD1L | 11/6/2 | 1 | $2.95 \times 10^{-3}$ | 11.39 | 0 | 0 |
| CHRNB2 | 11/7/1 | 1 | $2.95 \times 10^{-3}$ | 11.39 | 0 | 0 |
| CYP2A13 | 11/7/4 | 1 | $2.95 \times 10^{-3}$ | 11.39 | 0 | 0 |

**Table S11**: BipEx and BSC case-control counts of the top ten most significant genes in the BipEx BD primary gene-based analysis. Case and control columns denote the count of ultra-rare PTVs in the gene of interest with MAC ≤ 5 in the respective dataset.

| Gene | *P*-value |
| --- | --- |
| AKAP11 | $1.15 \times 10^{-5}$ |
| DOP1A | $2.22 \times 10^{-4}$ |
| SHANK1 | $8.19 \times 10^{-4}$ |
| TOPAZ1 | $1.56 \times 10^{-3}$ |
| ATP9A | $1.66 \times 10^{-3}$ |
| WWP1 | $6.52 \times 10^{-3}$ |
| HECTD2 | $6.91 \times 10^{-3}$ |
| PSAP | $1.41 \times 10^{-2}$ |
| RAP1GDS1 | $1.41 \times 10^{-2}$ |
| USP24 | $1.41 \times 10^{-2}$ |
| SPHKAP | $1.57 \times 10^{-2}$ |
| CACNA1B | $1.93 \times 10^{-2}$ |
| ANKFY1 | $1.95 \times 10^{-2}$ |
| SCN3A | $1.95 \times 10^{-2}$ |
| SMG7 | $1.95 \times 10^{-2}$ |
| DNAJC14 | $2.86 \times 10^{-2}$ |
| EXOC3 | $2.86 \times 10^{-2}$ |
| PHIP | $2.86 \times 10^{-2}$ |
| SBNO1 | $3.14 \times 10^{-2}$ |
| ZFYVE9 | $3.14 \times 10^{-2}$ |

**Table S12**: Top 20 genes with *p*LI ≥ 0.9 as measured by gene-based test *P*-value.

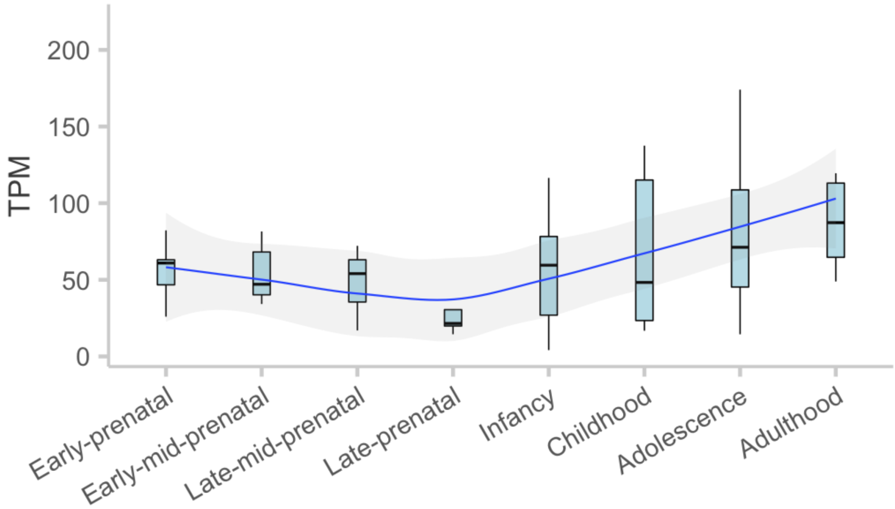


**Figure S16:** Temporal expression of AKAP11 in the human brain. Expression in four prenatal and four postnatal periods derived from whole-brain tissue in BrainSpan are displayed. The expression values plotted are in transcript-per-million (TPM). In each boxplot, the blue box encloses the interquartile range, with a horizontal line denoting the median. Best fit lines and confidence-intervals across the *x*-axis are overlaid.

#### References


1. American Psychiatric Association. Task Force on DSM-IV., *DSM-IV Sourcebook* (Amer Psychiatric Pub Incorporated, 1998).

2. A. Janca, T. B. Ustün, T. S. Early, N. Sartorius, The ICD-10 symptom checklist: a companion to the ICD-10 classification of mental and behavioural disorders. *Soc. Psychiatry Psychiatr. Epidemiol.* **28**, 239–242 (1993).

3. J. Wing, SCAN (Schedules for Clinical Assessment in Neuropsychiatry) and the PSE (Present State Examination) Tradition. *Mental Health Outcome Measures* (1996), pp. 123–130.

4. P. McGuffin, A. Farmer, I. Harvey, A polydiagnostic application of operational criteria in studies of psychotic illness. Development and reliability of the OPCRIT system. *Arch. Gen. Psychiatry*. **48**, 764–770 (1991).

5. T. J. C. Polderman, R. A. Hoekstra, A. A. E. Vinkhuyzen, P. F. Sullivan, S. van der Sluis, D. Posthuma, Attentional switching forms a genetic link between attention problems and autistic traits in adults. *Psychol. Med.* **43**, 1985–1996 (2013).

6. C. Keith Conners, D. Erhardt, E. P. Sparrow, *Conner's Adult ADHD Rating Scales: CAARS : Technical Manual* (1999).

7. T. M. Achenbach, *A Manual for the Young Adult Self-Report and Young Adult Behavior Checklist* (University of Vermont Department of Psychiatry, 1997).

8. S. Baron-Cohen, S. Wheelwright, R. Skinner, J. Martin, E. Clubley, The autism-spectrum quotient (AQ): evidence from Asperger syndrome/high-functioning autism, males and females, scientists and mathematicians. *J. Autism Dev. Disord.* **31**, 5–17 (2001).

9. M. B. First, R. L. Spitzer, J. B. Williams, M. Gibbon, *Structured Clinical Interview for DSM-IV Axis I Disorders (SCID-I), Clinician Version: Set of User's Guide, Administration Booklet, and Package of 5 Scoresheets* (Amer Psychiatric Pub Incorporated, 1997).

10. First, M.B., Spitzer, R.L., Gibbon, M., Williams, J.B.W., *Structured Clinical Interview for DSM-IV Axis I Disorders – Non-patient Edition. (SCID-I/NP)* (Biometrics Research, New York State Psychiatric Institute, New York., 1998).

11. P. P. Zandi, J. A. Badner, J. Steele, V. L. Willour, K. Miao, D. F. MacKinnon, F. M. Mondimore, B. Schweizer, M. G. McInnis, J. R. DePaulo Jr, E. Gershon, F. J. McMahon, J. B. Potash, Genome-wide linkage scan of 98 bipolar pedigrees and analysis of clinical covariates. *Mol. Psychiatry*. **12**, 630–639 (2007).

12. F. S. Goes, M. Pirooznia, J. S. Parla, M. Kramer, E. Ghiban, S. Mavruk, Y.-C. Chen, E. T. Monson, V. L. Willour, R. Karchin, M. Flickinger, A. E. Locke, S. E. Levy, L. J. Scott, M. Boehnke, E. Stahl, J. L. Moran, C. M. Hultman, M. Landén, S. M. Purcell, P. Sklar, P. P. Zandi, W. R. McCombie, J. B. Potash, Exome Sequencing of Familial Bipolar Disorder. *JAMA Psychiatry*. **73**, 590–597 (2016).

13. A. W. Charney, D. M. Ruderfer, E. A. Stahl, J. L. Moran, K. Chambert, R. A. Belliveau, L. Forty, K. Gordon-Smith, A. Di Florio, P. H. Lee, E. J. Bromet, P. F. Buckley, M. A. Escamilla, A. H. Fanous, L. J. Fochtmann, D. S. Lehrer, D. Malaspina, S. R. Marder, C. P. Morley, H. Nicolini, D. O. Perkins, J. J. Rakofsky, M. H. Rapaport, H. Medeiros, J. L. Sobell, E. K. Green, L. Backlund, S. E. Bergen, A. Juréus, M. Schalling, P. Lichtenstein, P. Roussos, J. A. Knowles, I. Jones, L. A. Jones, C. M. Hultman, R. H. Perlis, S. M. Purcell, S. A. McCarroll, C. N. Pato, M. T. Pato, N. Craddock, M. Landén, J. W. Smoller, P. Sklar, Evidence for genetic heterogeneity between clinical subtypes of bipolar

disorder. *Transl. Psychiatry*. **7**, e993 (2017).

14. C.-Y. Chen, P. H. Lee, V. M. Castro, J. Minnier, A. W. Charney, E. A. Stahl, D. M. Ruderfer, S. N. Murphy, V. Gainer, T. Cai, I. Jones, C. N. Pato, M. T. Pato, M. Landén, P. Sklar, R. H. Perlis, J. W. Smoller, Genetic validation of bipolar disorder identified by automated phenotyping using electronic health records. *Transl. Psychiatry*. **8**, 86 (2018).

15. V. M. Castro, J. Minnier, S. N. Murphy, I. Kohane, S. E. Churchill, V. Gainer, T. Cai, A. G. Hoffnagle, Y. Dai, S. Block, S. R. Weill, M. Nadal-Vicens, A. R. Pollastri, J. N. Rosenquist, S. Goryachev, D. Ongur, P. Sklar, R. H. Perlis, J. W. Smoller, International Cohort Collection for Bipolar Disorder Consortium, Validation of electronic health record phenotyping of bipolar disorder cases and controls. *Am. J. Psychiatry*. **172**, 363–372 (2015).

16. The ninth revision of the International Classification of Diseases: nosology and the specialist in community medicine. *Journal of Public Health* (1979), , doi:10.1093/oxfordjournals.pubmed.a043205.

17. V. N. Slee, The International Classification of Diseases: Ninth Revision (ICD-9). *Annals of Internal Medicine*. **88** (1978), p. 424.

18. Wellcome Trust Case Control Consortium, Genome-wide association study of 14,000 cases of seven common diseases and 3,000 shared controls. *Nature*. **447**, 661–678 (2007).

19. M. C. O'Donovan, N. Craddock, N. Norton, H. Williams, T. Peirce, V. Moskvina, I. Nikolov, M. Hamshere, L. Carroll, L. Georgieva, S. Dwyer, P. Holmans, J. L. Marchini, C. C. A. Spencer, B. Howie, H.-T. Leung, A. M. Hartmann, H.-J. Möller, D. W. Morris, Y. Shi, G. Feng, P. Hoffmann, P. Propping, C. Vasilescu, W. Maier, M. Rietschel, S. Zammit, J. Schumacher, E. M. Quinn, T. G. Schulze, N. M. Williams, I. Giegling, N. Iwata, M. Ikeda, A. Darvasi, S. Shifman, L. He, J. Duan, A. R. Sanders, D. F. Levinson, P. V. Gejman, S. Cichon, M. M. Nöthen, M. Gill, A. Corvin, D. Rujescu, G. Kirov, M. J. Owen, N. G. Buccola, B. J. Mowry, R. Freedman, F. Amin, D. W. Black, J. M. Silverman, W. F. Byerley, C. R. Cloninger, Molecular Genetics of Schizophrenia Collaboration, Identification of loci associated with schizophrenia by genome-wide association and follow-up. *Nat. Genet.* **40**, 1053–1055 (2008).

20. Schizophrenia Working Group of the Psychiatric Genomics Consortium, Biological insights from 108 schizophrenia-associated genetic loci. *Nature*. **511**, 421–427 (2014).

21. S. V. Parikh, S. R. LeBlanc, M. M. Ovanessian, Advancing bipolar disorder: key lessons from the Systematic Treatment Enhancement Program for Bipolar Disorder (STEP-BD). *Can. J. Psychiatry*. **55**, 136–143 (2010).

22. M. B. First, M. Gibbon, R. L. Spitzer, J. B. W. Williams, *Structured Clinical Interview for DSM-IV Axis II Personality Disorders: SCID-II* (Amer Psychiatric Pub Incorporated, 1997).

23. C. Sellgren, M. Landén, P. Lichtenstein, C. M. Hultman, N. Långström, Validity of bipolar disorder hospital discharge diagnoses: file review and multiple register linkage in Sweden. *Acta Psychiatr. Scand.* **124**, 447–453 (2011).

24. C. Almqvist, H.-O. Adami, P. W. Franks, L. Groop, E. Ingelsson, J. Kere, L. Lissner, J.-E. Litton, M. Maeurer, K. Michaëlsson, J. Palmgren, G. Pershagen, A. Ploner, P. F. Sullivan, G. Tybring, N. L. Pedersen, LifeGene—a large prospective population-based study of global relevance. *European Journal of Epidemiology*. **26** (2011), pp. 67–77.

25. Quality of life for patients diagnosed with bipolar disorder: Lifestyle and treatment. *Neurology, Psychiatry and Brain Research*. **34**, 34–40 (2019).

26. I. M. van Vliet, E. de Beurs, [The MINI-International Neuropsychiatric Interview. A brief structured

diagnostic psychiatric interview for DSM-IV en ICD-10 psychiatric disorders]. *Tijdschr. Psychiatr.* **49**, 393–397 (2007).

27. A. Frances, The Diagnostic Interview for Genetic Studies. *Archives of General Psychiatry*. **51** (1994), p. 863.

28. J. Endicott, R. L. Spitzer, A diagnostic interview: the schedule for affective disorders and schizophrenia. *Arch. Gen. Psychiatry*. **35**, 837–844 (1978).

29. S. N. Murphy, M. E. Mendis, D. A. Berkowitz, I. Kohane, H. C. Chueh, Integration of clinical and genetic data in the i2b2 architecture. *AMIA Annu. Symp. Proc.*, 1040 (2006).

30. R.-D. Stieglitz, A. Haug, E. Fähndrich, M. Rösler, W. Trabert, Comprehensive Psychopathological Assessment Based on the Association for Methodology and Documentation in Psychiatry (AMDP) System: Development, Methodological Foundation, Application in Clinical Routine, and Research. *Front. Psychiatry*. **8**, 45 (2017).

31. K. Kröger, A. Stang, J. Kondratieva, S. Moebus, E. Beck, A. Schmermund, S. Möhlenkamp, N. Dragano, J. Siegrist, K.-H. Jöckel, R. Erbel, Heinz Nixdorf Recall Study Group, Prevalence of peripheral arterial disease - results of the Heinz Nixdorf recall study. *Eur. J. Epidemiol.* **21**, 279–285 (2006).

32. G. A. Van der Auwera, M. O. Carneiro, C. Hartl, R. Poplin, G. Del Angel, A. Levy-Moonshine, T. Jordan, K. Shakir, D. Roazen, J. Thibault, E. Banks, K. V. Garimella, D. Altshuler, S. Gabriel, M. A. DePristo, From FastQ data to high confidence variant calls: the Genome Analysis Toolkit best practices pipeline. *Curr. Protoc. Bioinformatics*. **43**, 11.10.1–11.10.33 (2013).

33. M. A. DePristo, E. Banks, R. Poplin, K. V. Garimella, J. R. Maguire, C. Hartl, A. A. Philippakis, G. del Angel, M. A. Rivas, M. Hanna, A. McKenna, T. J. Fennell, A. M. Kernytsky, A. Y. Sivachenko, K. Cibulskis, S. B. Gabriel, D. Altshuler, M. J. Daly, A framework for variation discovery and genotyping using next-generation DNA sequencing data. *Nat. Genet.* **43**, 491–498 (2011).

34. H. Li, Toward better understanding of artifacts in variant calling from high-coverage samples. *Bioinformatics*. **30**, 2843–2851 (2014).

35. G. Jun, M. Flickinger, K. N. Hetrick, J. M. Romm, K. F. Doheny, G. R. Abecasis, M. Boehnke, H. M. Kang, Detecting and estimating contamination of human DNA samples in sequencing and array-based genotype data. *Am. J. Hum. Genet.* **91**, 839–848 (2012).

36. S. Purcell, B. Neale, K. Todd-Brown, L. Thomas, M. A. R. Ferreira, D. Bender, J. Maller, P. Sklar, P. I. W. de Bakker, M. J. Daly, P. C. Sham, PLINK: a tool set for whole-genome association and population-based linkage analyses. *Am. J. Hum. Genet.* **81**, 559–575 (2007).

37. C. C. Chang, C. C. Chow, L. C. Tellier, S. Vattikuti, S. M. Purcell, J. J. Lee, Second-generation PLINK: rising to the challenge of larger and richer datasets. *Gigascience*. **4**, 7 (2015).

38. T. 1000 G. P. Consortium, The 1000 Genomes Project Consortium, A global reference for human genetic variation. *Nature*. **526** (2015), pp. 68–74.

39. W. McLaren, L. Gil, S. E. Hunt, H. S. Riat, G. R. S. Ritchie, A. Thormann, P. Flicek, F. Cunningham, The Ensembl Variant Effect Predictor. *Genome Biol.* **17**, 122 (2016).

40. P. C. Ng, S. Henikoff, SIFT: Predicting amino acid changes that affect protein function. *Nucleic Acids Res.* **31**, 3812–3814 (2003).

41. I. A. Adzhubei, S. Schmidt, L. Peshkin, V. E. Ramensky, A. Gerasimova, P. Bork, A. S. Kondrashov,

S. R. Sunyaev, A method and server for predicting damaging missense mutations. *Nat. Methods*. **7**, 248–249 (2010).

42. K. J. Karczewski, L. C. Francioli, G. Tiao, B. B. Cummings, J. Alföldi, Q. Wang, R. L. Collins, K. M. Laricchia, A. Ganna, D. P. Birnbaum, L. D. Gauthier, H. Brand, M. Solomonson, N. A. Watts, D. Rhodes, M. Singer-Berk, E. M. England, E. G. Seaby, J. A. Kosmicki, R. K. Walters, K. Tashman, Y. Farjoun, E. Banks, T. Poterba, A. Wang, C. Seed, N. Whiffin, J. X. Chong, K. E. Samocha, E. Pierce-Hoffman, Z. Zappala, A. H. O'Donnell-Luria, E. V. Minikel, B. Weisburd, M. Lek, J. S. Ware, C. Vittal, I. M. Armean, L. Bergelson, K. Cibulskis, K. M. Connolly, M. Covarrubias, S. Donnelly, S. Ferriera, S. Gabriel, J. Gentry, N. Gupta, T. Jeandet, D. Kaplan, C. Llanwarne, R. Munshi, S. Novod, N. Petrillo, D. Roazen, V. Ruano-Rubio, A. Saltzman, M. Schleicher, J. Soto, K. Tibbetts, C. Tolonen, G. Wade, M. E. Talkowski, Genome Aggregation Database Consortium, B. M. Neale, M. J. Daly, D. G. MacArthur, The mutational constraint spectrum quantified from variation in 141,456 humans. *Nature*. **581**, 434–443 (2020).

43. K. E. Samocha, J. A. Kosmicki, K. J. Karczewski, A. H. O'Donnell-Luria, E. Pierce-Hoffman, D. G. MacArthur, B. M. Neale, M. J. Daly, Regional missense constraint improves variant deleteriousness prediction, , doi:10.1101/148353.

44. P. Rentzsch, D. Witten, G. M. Cooper, J. Shendure, M. Kircher, CADD: predicting the deleteriousness of variants throughout the human genome. *Nucleic Acids Res.* **47**, D886–D894 (2019).

45. G. Genovese, M. Fromer, E. A. Stahl, D. M. Ruderfer, K. Chambert, M. Landén, J. L. Moran, S. M. Purcell, P. Sklar, P. F. Sullivan, C. M. Hultman, S. A. McCarroll, Increased burden of ultra-rare protein-altering variants among 4,877 individuals with schizophrenia. *Nature Neuroscience*. **19** (2016), pp. 1433–1441.

46. T. Singh, J. T. R. Walters, M. Johnstone, D. Curtis, J. Suvisaari, M. Torniainen, E. Rees, C. Iyegbe, D. Blackwood, A. M. McIntosh, G. Kirov, D. Geschwind, R. M. Murray, M. Di Forti, E. Bramon, M. Gandal, C. M. Hultman, P. Sklar, INTERVAL Study, UK10K Consortium, A. Palotie, P. F. Sullivan, M. C. O'Donovan, M. J. Owen, J. C. Barrett, The contribution of rare variants to risk of schizophrenia in individuals with and without intellectual disability. *Nat. Genet.* **49**, 1167–1173 (2017).

47. S. Murphy, S. Churchill, L. Bry, H. Chueh, S. Weiss, R. Lazarus, Q. Zeng, A. Dubey, V. Gainer, M. Mendis, J. Glaser, I. Kohane, Instrumenting the health care enterprise for discovery research in the genomic era. *Genome Res.* **19**, 1675–1681 (2009).

48. H. K. Finucane, Y. A. Reshef, V. Anttila, K. Slowikowski, A. Gusev, A. Byrnes, S. Gazal, P.-R. Loh, C. Lareau, N. Shoresh, G. Genovese, A. Saunders, E. Macosko, S. Pollack, Brainstorm Consortium, J. R. B. Perry, J. D. Buenrostro, B. E. Bernstein, S. Raychaudhuri, S. McCarroll, B. M. Neale, A. L. Price, Heritability enrichment of specifically expressed genes identifies disease-relevant tissues and cell types. *Nat. Genet.* **50**, 621–629 (2018).

49. T. Singh, T. Poterba, D. Curtis, H. Akil, M. Al Eissa, J. D. Barchas, N. Bass, T. B. Bigdeli, G. Breen, E. J. Bromet, P. F. Buckley, W. E. Bunney, J. Bybjerg-Grauholm, W. F. Byerley, S. B. Chapman, W. J. Chen, C. Churchhouse, N. Craddock, C. Curtis, C. M. Cusick, L. DeLisi, S. Dodge, M. A. Escamilla, S. Eskelinen, A. H. Fanous, S. V. Faraone, A. Fiorentino, L. Francioli, S. B. Gabriel, D. Gage, S. A. Gagliano Taliun, A. Ganna, G. Genovese, D. C. Glahn, J. Grove, M.-H. Hall, E. Hamalainen, H. O. Heyne, M. Holi, D. M. Hougaard, D. P. Howrigan, H. Huang, H.-G. Hwu, R. S. Kahn, H. M. Kang, K. Karczewski, G. Kirov, J. A. Knowles, F. S. Lee, D. S. Lehrer, F. Lescai, D. Malaspina, S. R. Marder, S. A. McCarroll, H. Medeiros, L. Milani, C. P. Morley, D. W. Morris, P. B. Mortensen, R. M. Myers, M. Nordentoft, N. L. O'Brien, A. M. Olivares, D. Ongur, W. H. Ouwehand, D. S. Palmer, T. Paunio, D. Quested, M. H. Rapaport, E. Rees, B. Rollins, F. Kyle Satterstrom, A. Schatzberg, E. Scolnick, L. Scott, S. I. Sharp, P. Sklar, J. W. Smoller, J. I. Sobell, M. Solomonson, C. R. Stevens, J. Suvisaari, G. Tiao, S. J. Watson, N. A. Watts, D. H. Blackwood, A. Borglum, B. M. Cohen, A. P. Corvin, T. Esko, N.

B. Freimer, S. J. Glatt, C. M. Hultman, A. McQuillin, A. Palotie, C. N. Pato, M. T. Pato, A. E. Pulver, D. St. Clair, M. T. Tsuang, M. P. Vawter, J. T. Walters, T. Werge, R. A. Ophoff, P. F. Sullivan, M. J. Owen, M. Boehnke, M. O'Donovan, B. M. Neale, M. J. Daly, Exome sequencing identifies rare coding variants in 10 genes which confer substantial risk for schizophrenia. *medRxiv*, 2020.09.18.20192815 (2020).

50. G. Jun, M. K. Wing, G. R. Abecasis, H. M. Kang, An efficient and scalable analysis framework for variant extraction and refinement from population-scale DNA sequence data. *Genome Res.* **25**, 918–925 (2015).
